# Contraceptive use and pregnancy planning in Britain during the first year of the COVID-19 pandemic: findings from a large, quasi-representative survey (Natsal-COVID)

**DOI:** 10.1101/2022.10.14.22281078

**Authors:** Andrew Baxter, Rebecca S. Geary, Emily Dema, Raquel Bosó Pérez, Julie Riddell, Malachi Willis, Anne Conolly, Laura Oakley, Andrew Copas, Jo Gibbs, Chris Bonell, Pam Sonnenberg, Catherine H. Mercer, Soazig Clifton, Nigel Field, Kirstin Mitchell

**Affiliations:** MRC/CSO Social and Public Health Sciences Unit, University of Glasgow, Glasgow, UK; Institute of Population Health, University of Liverpool, Liverpool, UK; Institute for Global Health, University College London, London, UK; NatCen Social Research, London, UK; London School of Hygiene and Tropical Medicine, London, UK; Centre for Fertility and Health, Norwegian Institute of Public Health, Oslo, Norway

## Abstract

**Background:** Reproductive health services were significantly disrupted during the COVID-19 pandemic in Britain. We investigated contraception-related health inequalities in the first year of the pandemic.

**Methods:** Natsal-COVID Wave 2 surveyed 6,658 adults aged 18–59 between March–April 2021, using quotas and weighting to achieve quasi-representativeness. Our analysis included sexually active participants aged 18–44, described as female at birth. We analysed contraception use and switching, contraceptive service access, and pregnancy plannedness in the year from March 2020.

**Findings:** Amongst all participants (n=1,488), 14.3% (12.5%-16.3%) reported switching or stopping contraception due to the pandemic. Of participants at risk of unplanned pregnancy (n=1,169), 54.1% (51.0%-57.1%) reported routinely using effective contraception in the past year. 3.2% (2.0%-5.1%) of those using effective methods pre-pandemic switched to less effective methods, while 3.8% (2.5%-5.9%) stopped. Stopping/switching was more likely amongst participants of younger age, non-white ethnicity, and lower social grade. 29.3% of at-risk participants (26.9%-31.8%) reported trying to access contraceptive services; of whom 16.4% (13.0%-20.4%) reported their needs went unmet. Unmet need was associated with younger age, diverse sexual identities and anxiety symptoms. Of 199 pregnancies, 6.6% (3.9%-11.1%) were scored as ‘unplanned’; less planning was associated with younger age, lower social grade and unemployment.

**Interpretation:** Although many participants reported accessing contraceptive services during the pandemic, one-in-six of these reported an unmet need. Inequalities in unmet need and risk of unplanned pregnancy – related to age, ethnicity, social disadvantage and mental health – potentially exacerbated existing reproductive health inequalities. These should be addressed in the post-pandemic period and beyond.

**Funding:** Wellcome Trust, The Economic and Social Research Council, The National Institute for Health Research, Medical Research Council/Chief Scientist Office Social and Public Health Sciences Unit, and UCL Coronavirus Response Fund.

**Key messages:** *What is already known on this topic:*

- The COVID-19 pandemic likely impacted reproductive outcomes in diverse ways; such impacts may have been unequally distributed.
- Previous studies reported adaptations to health service delivery and difficulties experienced in accessing reproductive health services, with switching and stopping of contraceptive methods and potentially greater risk of unplanned pregnancy.

*What this study adds:*

- We examined differences in contraceptive use and pregnancy planning in a sample of women, trans and non-binary people able to become pregnant who were quasi-representative of the British general population.
- We found that key markers of inequality and vulnerability, related to age, ethnicity, social disadvantage and mental health, were associated with increased contraceptive method switching, unmet need of contraceptive services and less-planned pregnancies.

*How this study might affect research, practice or policy:*

- Ongoing efforts to ease the health impacts of the pandemic should aim to improve equality of access to contraceptive services.

## Introduction

The COVID-19 pandemic prompted rapid adjustment to health services, including the suspension or reduction of face-to-face consultations, increased remote provision, and rearranged appointments due to staff unavailability.^1,2^ Adjustments to contraceptive services included recommendation of methods not requiring face-to-face consultations (e.g. the progestogen-only pill), amended guidance on off-label extended use of some long-acting reversible contraceptives (LARCs), and streamlined remote repeat prescribing of the combined contraceptive pill.^3^

Although aiming for equitable access,^4,5^ rapid adaptions during the pandemic had the potential to exacerbate inequalities, particularly if these required digital access and literacy.^6,7^ Service users might also interpret adaptations as de-prioritising contraceptive services,^8^ and we know that some patients self-censored their needs or were anxious about COVID-19 risk if accessing services in person.^9^ Overall, people in the UK and globally struggled to access contraception during lockdowns,^1,10–14^ and prescribing data for the UK showed substantial drops in LARCs fitted in 2020 versus 2019.^15^ Several studies suggest that young people were disproportionately affected by service closures.^10,16,17^

However, the pandemic’s effect on contraception and service use remains poorly understood. Previous studies have indicated difficulties accessing contraception, alongside changing sexual risk behaviours, however these often used small convenience samples in the early stages of the pandemic.^8,14,17,18^ The Natsal-COVID study, a large national survey of sexual and reproductive health, was set up to address gaps in representativeness of studies and a lack of detailed information about the ongoing effects of the pandemic. Wave 1 findings (conducted four months after the first UK national lockdown) suggested young women were most likely to switch contraceptive and face barriers to sexual health service access.^11^ We have also reported Wave 2 findings that one-in-10 participants described as female at birth had stopped or switched contraceptive method in the year after the first lockdown.^19^ In this study, we investigate inequalities in reproductive health service access, contraceptive method switching due to the pandemic and pregnancy ‘plannedness’ during the first year of the pandemic amongst women, and trans and non-binary people who can become pregnant.

## Methods

### Study design and participants

Natsal-COVID Wave 2 is a quasi-representative web panel survey of sexual and reproductive health (SRH) in Britain. Wave 2 survey data were collected between 27/03/21-26/04/21.^20,21^ Participants aged 18-59 answered an online questionnaire administered by Ipsos (median length 13-minutes). Participants were asked about SRH and service use in the period before the first UK lockdown in March 2020 and in the past year. Participants were asked their gender identity, sex described at birth, sexual orientation, ethnicity, education, general health and disability, mental health and alcohol consumption. The full questionnaire is available online.^22^

We used sampling target quotas set by gender, age, region and social grade and weighting based on gender, age, region, social grade, ethnicity and sexual orientation to achieve a quasi-representative sample of the British general population.^20,21^

### Statistical analysis

The Wave 2 sample (n=6,658) comprised 2,098 recontacted Wave 1 participants and 4,560 new participants aged 18-59. The latter included a boost of 500 people aged 18-29, ensuring an overall sample of 2,000 participants in this age group, who are often at greater risk of adverse SRH outcomes.^20^

To examine the impacts of the covid pandemic on pregnancy planning, we analysed prevalence and plannedness of pregnancy among participants aged 18–44 who were described as female at birth and reported any sexual contact with a man since the start of the first UK lockdown (23/03/2020). We analysed contraceptive method use and service access among a sub-sample of those at risk of unplanned pregnancy that we defined by excluding those currently pregnant, currently trying to conceive or not able to get pregnant.

To measure inequalities, we used educational attainment by highest academic qualification reported, and social grade based on occupation. Participants were classified as having symptoms of depression or anxiety if scoring <u>>3</u> on the two-item patient health questionnaire (PHQ-2) or generalised anxiety disorder (GAD-2) screening tools respectively.^23,24^

We categorised contraceptive methods by their effectiveness in preventing pregnancy based on typical-use failure rates (Box 1).^25,26^ We analysed emergency contraceptive (EC) use separately, assuming that changes in motivation and access would have affected use of planned methods and EC differently. Participants using another method in addition to EC were classified by the effectiveness of the non-emergency method; participants who only used EC were classified as using ‘no method’ for the purposes of prophylactic method-use comparisons. Unmet need for contraceptive services was defined as reporting trying but being unable to use contraceptive services. ‘Plannedness’ of pregnancies in the past year was estimated using the London Measure of Unplanned Pregnancy (LMUP, 2020 version),^27,28^ comprising six questions on contraceptive use, timing of motherhood, intention to become pregnant, desire for a baby, discussion with a partner and pre-conception health preparations.^29^ Each item is scored 0–2 (summing to a total, range=0–12), with each point representing an increase in pregnancy ‘plannedness’. Scores of 0–3 were categorised as ‘unplanned’, 4–9 as ‘ambivalent’ and above 9 as ‘planned’. Full definitions for outcome variables and the denominators used in each analysis are given in Supplementary Table 1.

We used complex survey analysis functions in Stata (version 17.0). Figures were constructed in R (version 4.2.1).^30^ Weighted estimates are presented with weighted and unweighted denominators and unweighted numerators. We used the survey-equivalent chi-square statistic to determine whether there was statistically significant variation in the reported contraceptive method used since the start of the first lockdown, or in the switching of contraceptive methods, by sociodemographic and behavioural factors. We compared odds of using EC pre- and post-lockdown, using a conditional logistic regression model to account for intra-person clustering. We used logistic regression to calculate age-adjusted odds ratios (aOR) to investigate how use of, and unmet need for, contraceptive services varied by sociodemographic and behavioural factors. We used linear regression with robust standard errors to investigate differences in mean ‘plannedness’ of pregnancy scores and logistic regression to estimate the odds of an ‘unplanned pregnancy’, adjusting for age, across sociodemographic and behavioural factors. Proportions of missing demographic variables were relatively low, ranging from 0% to 1.3%; all comparisons were restricted to complete cases across relevant variables.

### Role of the funding source

The funders had no role in the study design, collection, analysis and interpretation of data, or writing.

### Ethical approval

Natsal-COVID was approved by ethics committees at the University of Glasgow (20019174) and the London School of Hygiene and Tropical Medicine (22565). Participants provided informed consent electronically at the start of the survey.

## Results

Of 6,658 participants in Natsal-COVID Wave 2, 1,488 were aged 18–44, described as female at birth and reported sexual contact with a man in the past year. Of these, most identified as ‘female’ (weighted proportion: 99.2%), two described themselves as ‘male’ and 10 described themselves ‘in another way’. Most participants were white (86.7%), married or in a steady cohabiting relationship (70.8%) and identified as heterosexual (96.7%).

Of participants who provided information about contraceptive use, 78.0% (n=1,169) were deemed to be at risk of unplanned pregnancy (excluding 246 who were currently pregnant, currently trying to conceive or not able to get pregnant). Just over half of participants at risk reported using a more effective contraceptive method as their usual or only contraceptive method in the past year (54.1%; Table 2). This was lowest among participants aged 18–24 (45.7% (38.7%-53.0%)), and considerably lower for participants from Black (27.6% (15.1%-45.0%)), Asian or Asian-British ethnic backgrounds (25.9% (15.8%-39.4%)) or from a mixed or multiple or other ethnic background (27.5% (15.2%-44.7%)) than participants from White ethnic backgrounds (58.1% (54.7%-61.3%)). Those with at least one marker of lower socioeconomic status (working in less skilled occupations, receiving state benefit or unemployed at the time of survey) were less likely to report using a more effective method as their only/usual contraceptive method (D/E social grade: 42.9% (36.7%-49.2%), vs 60.9% (56.6%-65.0%) of C1/C2 social grade). However, economic factors potentially related to the COVID-19 pandemic were not associated with effective contraception use (becoming unemployed: p=0.20; or furloughed: p=0.90; Table 2). Among those at risk of pregnancy, reported EC use was higher in the year preceding the pandemic (reported by 3.4% (2.4%-4.9%)) than in the year from the start of the first lockdown (1.9% (1.1%-3.2%); OR: 0.30 (0.12-0.75); data not shown).

**Table 1.** Socio-demographic characteristics of sexually active participants aged 18-44 years, described as female at birth who reported sex with a man in the past year.

| Age group | 18-24 | 25-29 | 30-34 | 35-44 | All ages |
| --- | --- | --- | --- | --- | --- |
| **Total** |  |  |  |  |  |
| Distribution across age categories (% (95% CI)) | 16.7<br>(15.3, 18.1) | 25.3<br>(23.7, 27.1) | 21.3<br>(19.7, 22.9) | 36.7<br>(34.9, 38.6) | 100.0% |
| Denominators (weighted, unweighted) | 213, 265 | 324, 397 | 272, 320 | 470, 506 | 1279, 1488 |
| **Distributions within age categories** |  |  |  |  |  |
|  | % (95% CI) | % (95% CI) | % (95% CI) | % (95% CI) | % (95% CI) |
| **Ethnicity** |  |  |  |  |  |
| White | 79.0<br>(72.9, 84.0) | 86.0<br>(81.8, 89.4) | 86.4<br>(81.8, 90.0) | 90.8<br>(87.8, 93.1) | 86.7%<br>(84.7, 88.5) |
| Black or Black African or Black Caribbean or Black British | 9.4 (6.1, 14.3) | 5.4 (3.4, 8.5) | 1.6 (0.6, 4.1) | 0.8 (0.3, 2.2) | 3.6% (2.7, 4.7) |
| Asian or Asian British | 8.0 (5.0, 12.5) | 4.2 (2.5, 7.1) | 7.7 (5.1, 11.6) | 6.0 (4.2, 8.6) | 6.2% (5.0, 7.7) |
| Mixed or multiple or other ethnic groups | 3.6 (1.8, 7.2) | 4.4 (2.6, 7.2) | 4.2 (2.4, 7.4) | 2.4 (1.3, 4.2) | 3.5% (2.6, 4.6) |
| Missing: n=10 (0.7%) |  |  |  |  |  |
| **Self-described sexual identity** |  |  |  |  |  |
| Heterosexual or Straight | 92.4<br>(88.0, 95.3) | 96.1<br>(93.3, 97.7) | 97.7<br>(95.0, 98.9) | 98.5<br>(96.9, 99.3) | 96.7%<br>(95.5, 97.5) |
| Lesbian, Gay, Bisexual or Other | 7.6 (4.7, 12.0) | 3.9 (2.3, 6.7) | 2.3 (1.1, 5.0) | 1.5 (0.7, 3.1) | 3.3% (2.5, 4.5) |
| Missing: n=12 (0.8%) |  |  |  |  |  |
| **Social Grade** |  |  |  |  |  |
| A/B - Higher/intermediate managerial, administrative and professional | 17.6<br>(13.0, 23.3) | 29.8<br>(25.1, 35.0) | 23.7<br>(19.0, 29.2) | 19.6<br>(16.2, 23.4) | 22.7%<br>(20.5, 25.1) |
| C1/C2 - Supervisory, clerical and junior managerial, administrative and professional, skilled manual workers | 39.3<br>(32.9, 46.0) | 49.1<br>(43.7, 54.6) | 50.3<br>(44.3, 56.2) | 61.4<br>(56.9, 65.7) | 52.3%<br>(49.5, 55.0) |
| D/E - Semi-skilled and unskilled manual, casual, lowest grade and unemployed | 43.1<br>(36.6, 49.9) | 21.1<br>(16.9, 25.9) | 26.0<br>(21.1, 31.6) | 19.0<br>(15.7, 22.8) | 25.0%<br>(22.7, 27.4) |
| Missing: n=0 (0%) |  |  |  |  |  |
| **Education level** |  |  |  |  |  |
| Degree | 28.3<br>(22.6, 34.7) | 58.5<br>(53.1, 63.8) | 59.0<br>(53.1, 64.7) | 54.8<br>(50.2, 59.2) | 52.2%<br>(49.5, 55.0) |
| Below degree | 65.1<br>(58.5, 71.3) | 36.7<br>(31.6, 42.1) | 37.2<br>(31.6, 43.1) | 43.4<br>(39.0, 47.9) | 44.0%<br>(41.3, 46.8) |
| No qualifications | 6.6 (3.9, 10.9) | 4.7 (2.9, 7.7) | 3.8 (2.1, 6.8) | 1.8 (0.9, 3.5) | 3.8% (2.8, 4.9) |
| Missing: n=0 (0%) |  |  |  |  |  |
| **Living together – relationship but not living together – Single** |  |  |  |  |  |
| Married/steady and living together | 34.4 (28.3, 41.0) | 71.0 (65.8, 75.7) | 78.5 (73.2, 83.0) | 82.2 (78.4, 85.4) | 70.6% (68.0, 73.0) |
| Married/steady NOT living together | 29.2 (23.5, 35.7) | 11.7 (8.6, 15.7) | 7.7 (5.1, 11.6) | 7.3 (5.3, 10.0) | 12.1% (10.4, 14.0) |
| Casual, new, >1, at end or other | 18.5 (13.8, 24.3) | 7.7 (5.3, 11.2) | 4.2 (2.4, 7.4) | 3.8 (2.4, 6.0) | 7.3% (6.0, 8.9) |
| Single | 17.9 (13.3, 23.7) | 9.5 (6.8, 13.3) | 9.6 (6.6, 13.7) | 6.7 (4.8, 9.4) | 9.9% (8.4, 11.7) |
| Missing: n=1 (0.1%) |  |  |  |  |  |
| **Been furloughed under the Coronavirus Job Retention Scheme** |  |  |  |  |  |
| No | 82.8<br>(77.0, 87.3) | 85.5<br>(81.2, 88.9) | 81.0<br>(75.9, 85.3) | 83.1<br>(79.4, 86.2) | 83.2%<br>(81.0, 85.2) |
| Yes | 17.2<br>(12.7, 23.0) | 14.5<br>(11.1, 18.8) | 19.0<br>(14.7, 24.1) | 16.9<br>(13.8, 20.6) | 16.8%<br>(14.8, 19.0) |
| Missing: n=12 (0.8%) |  |  |  |  |  |
| **Became unemployed** |  |  |  |  |  |
| No | 86.2<br>(80.8, 90.3) | 92.4<br>(89.0, 94.9) | 91.9<br>(88.0, 94.6) | 91.9<br>(89.0, 94.1) | 91.1%<br>(89.4, 92.5) |
| Yes | 13.8<br>(9.7, 19.2) | 7.6 (5.1, 11.0) | 8.1 (5.4, 12.0) | 8.1 (5.9, 11.0) | 8.9%<br>(7.5, 10.6) |
| Missing: n=12 (0.8%) |  |  |  |  |  |
| **Number of days drinking in past week** |  |  |  |  |  |
| 0 days | 38.0<br>(31.7, 44.7) | 40.1<br>(34.9, 45.5) | 46.0<br>(40.2, 52.0) | 45.7<br>(41.2, 50.2) | 43.0%<br>(40.3, 45.8) |
| 1-2 days | 41.0<br>(34.6, 47.8) | 40.9<br>(35.6, 46.4) | 39.5<br>(33.9, 45.5) | 35.3<br>(31.1, 39.7) | 38.6%<br>(35.9, 41.3) |
| 3-4 days | 17.3<br>(12.8, 23.0) | 13.1<br>(9.8, 17.3) | 9.5 (6.5, 13.7) | 11.3<br>(8.7, 14.5) | 12.4%<br>(10.7, 14.3) |
| 5-7 days | 3.7 (1.8, 7.2) | 5.9 (3.8, 9.1) | 4.9 (2.9, 8.2) | 7.8 (5.7, 10.6) | 6.0% (4.8, 7.5) |
| Missing: n=2 (0.1%) |  |  |  |  |  |
| **Drinking habits compared to pre Covid-19 outbreak** |  |  |  |  |  |
| Less these days | 36.9<br>(30.6, 43.6) | 30.9<br>(26.1, 36.2) | 28.8<br>(23.7, 34.6) | 25.8<br>(22.0, 30.0) | 29.6%<br>(27.1, 32.2) |
| About the same | 40.7<br>(34.2, 47.4) | 51.6<br>(46.1, 57.1) | 57.2<br>(51.1, 63.0) | 56.9<br>(52.4, 61.4) | 52.9%<br>(50.1, 55.6) |
| More these days | 22.4<br>(17.3, 28.6) | 17.5<br>(13.7, 22.0) | 14.0<br>(10.3, 18.7) | 17.3<br>(14.1, 21.0) | 17.5%<br>(15.5, 19.7) |
| Missing: n=19 (1.3%) |  |  |  |  |  |
| **Current smoker** |  |  |  |  |  |
| No | 75.9<br>(69.7, 81.2) | 73.4<br>(68.3, 77.9) | 79.8<br>(74.6, 84.2) | 82.3<br>(78.5, 85.5) | 78.5%<br>(76.1, 80.6) |
| Yes | 24.1<br>(18.8, 30.3) | 26.6<br>(22.1, 31.7) | 20.2<br>(15.8, 25.4) | 17.7<br>(14.5, 21.5) | 21.5%<br>(19.4, 23.9) |
| Missing: n=2 (0.1%) |  |  |  |  |  |
| **Symptoms of depression (PHQ2 score)** |  |  |  |  |  |
| No symptoms of depression (0-2) | 47.8<br>(41.0, 54.6) | 63.1<br>(57.6, 68.2) | 67.2<br>(61.3, 72.5) | 74.6<br>(70.4, 78.3) | 65.7%<br>(63.1, 68.3) |
| Symptoms of depression (3-6) | 52.2<br>(45.4, 59.0) | 36.9<br>(31.8, 42.4) | 32.8<br>(27.5, 38.7) | 25.4<br>(21.7, 29.6) | 34.3%<br>(31.7, 36.9) |
| Missing: n=15 (1.0%) |  |  |  |  |  |
| **Symptoms of anxiety (GAD2 score)** |  |  |  |  |  |
| No symptoms of anxiety (0-2) | 46.4<br>(39.7, 53.3) | 58.3<br>(52.8, 63.5) | 63.7<br>(57.8, 69.2) | 69.4<br>(65.0, 73.4) | 61.6%<br>(58.9, 64.2) |
| Symptoms of anxiety (3-6) | 53.6<br>(46.7, 60.3) | 41.7<br>(36.5, 47.2) | 36.3<br>(30.8, 42.2) | 30.6<br>(26.6, 35.0) | 38.4%<br>(35.8, 41.1) |
| Missing: n=8 (0.5%) |  |  |  |  |  |

\* In a 'casual' relationship, in a 'new' relationship, in more than one relationship, recently ended a relationship or 'other' relationship status

**Table 2.**
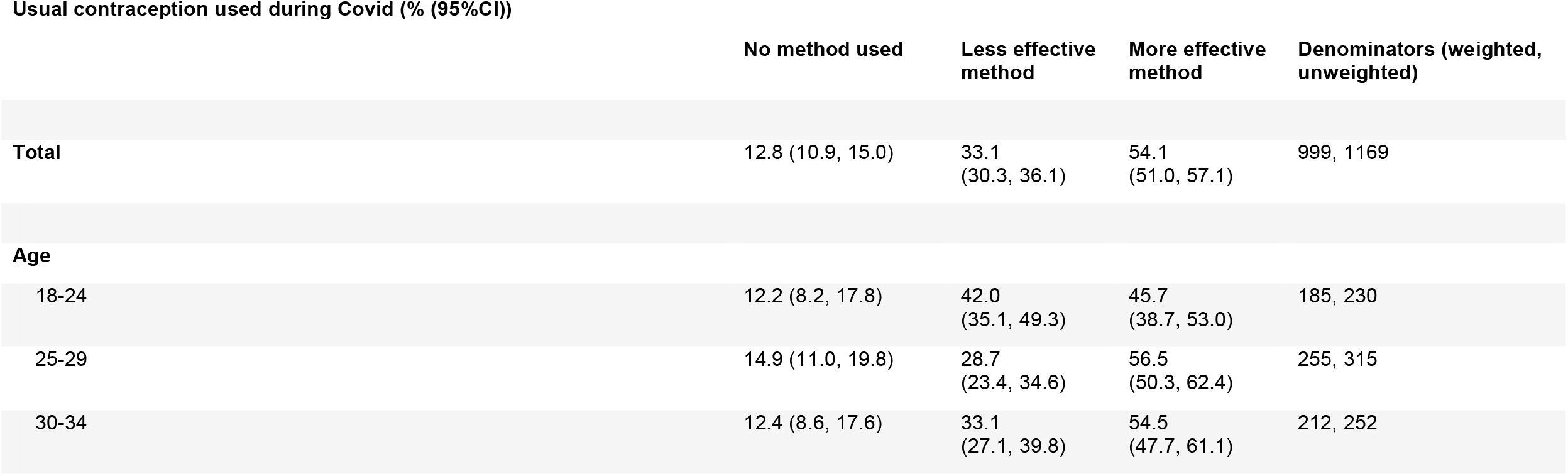
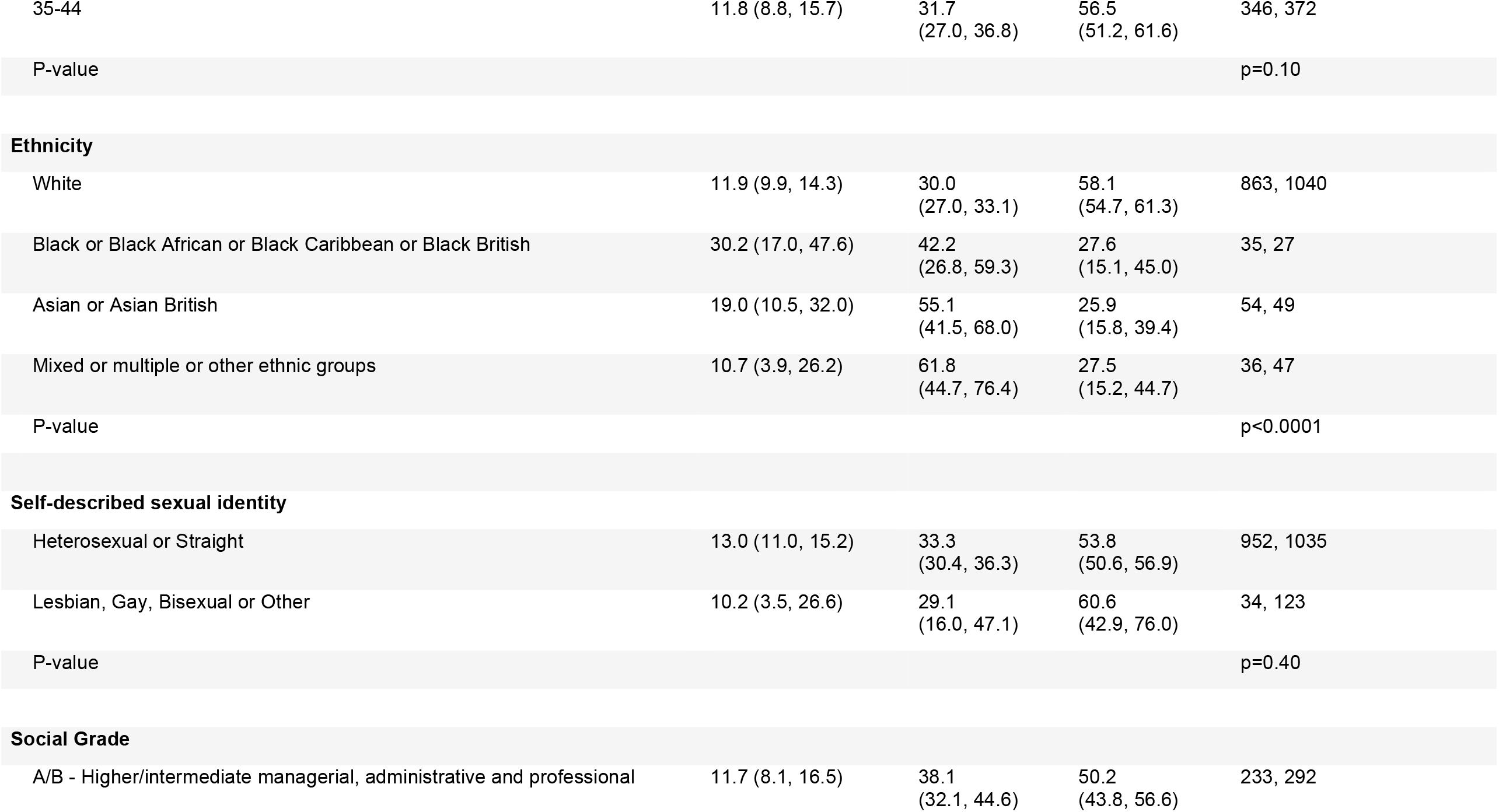
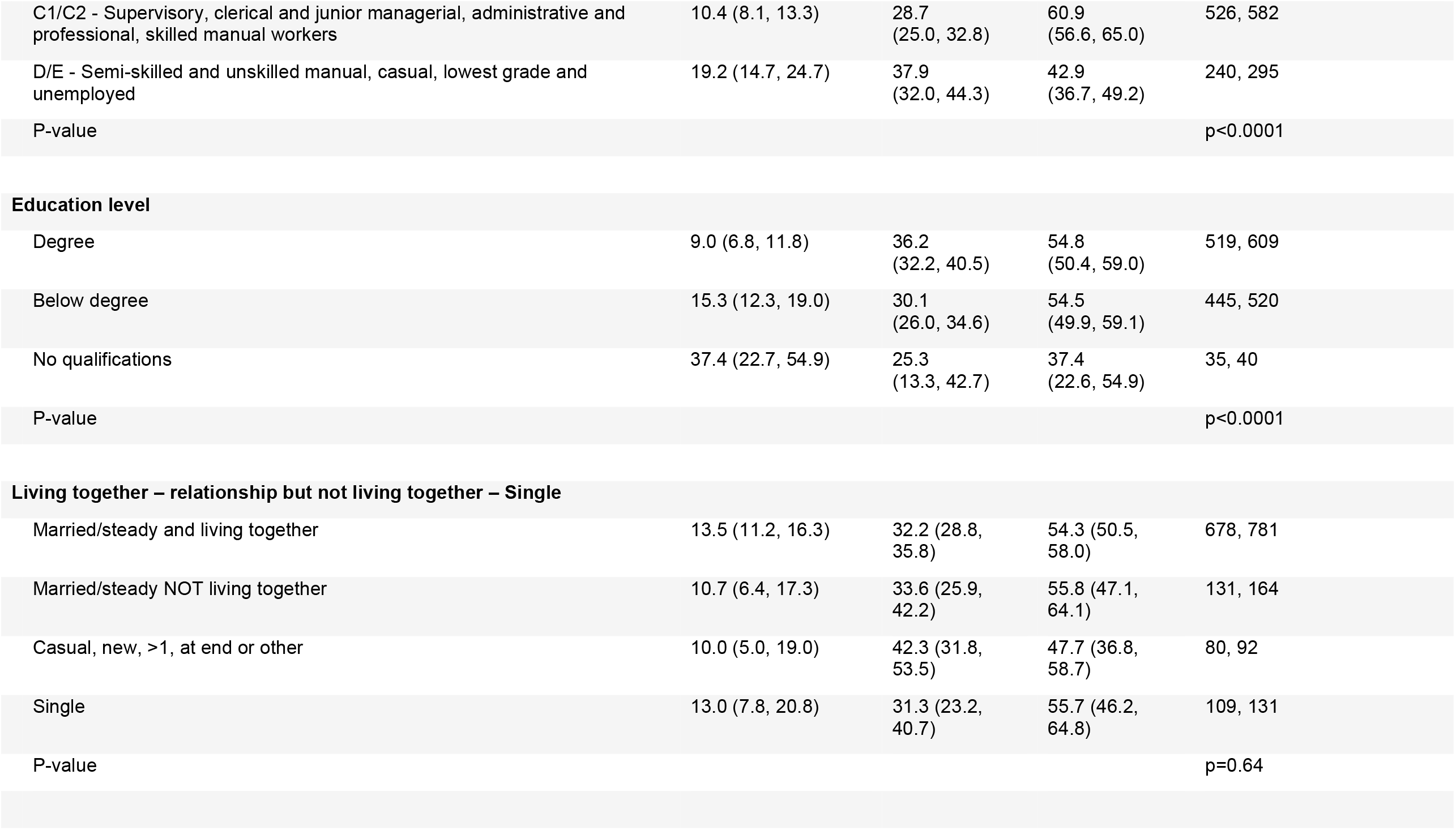
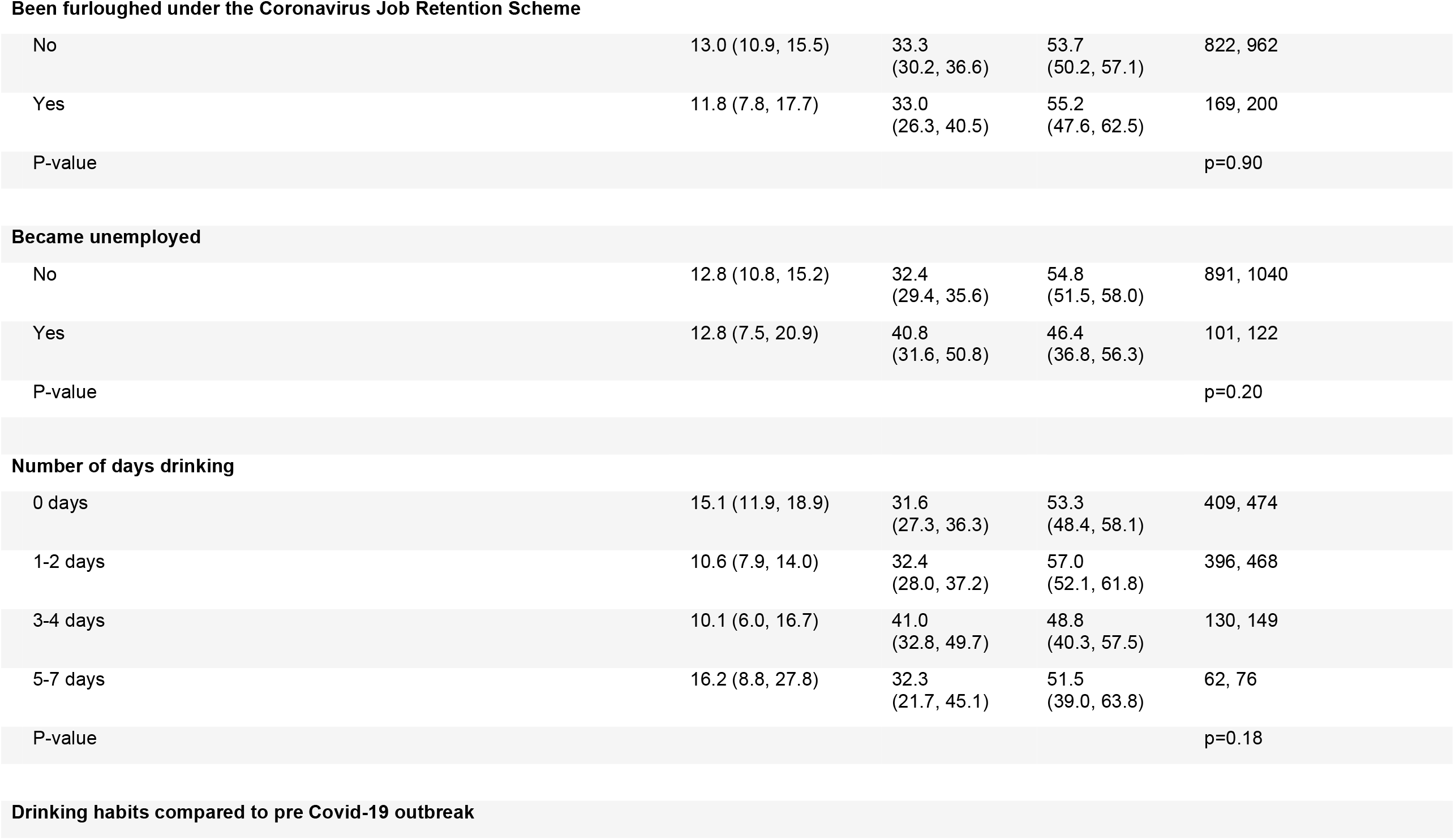
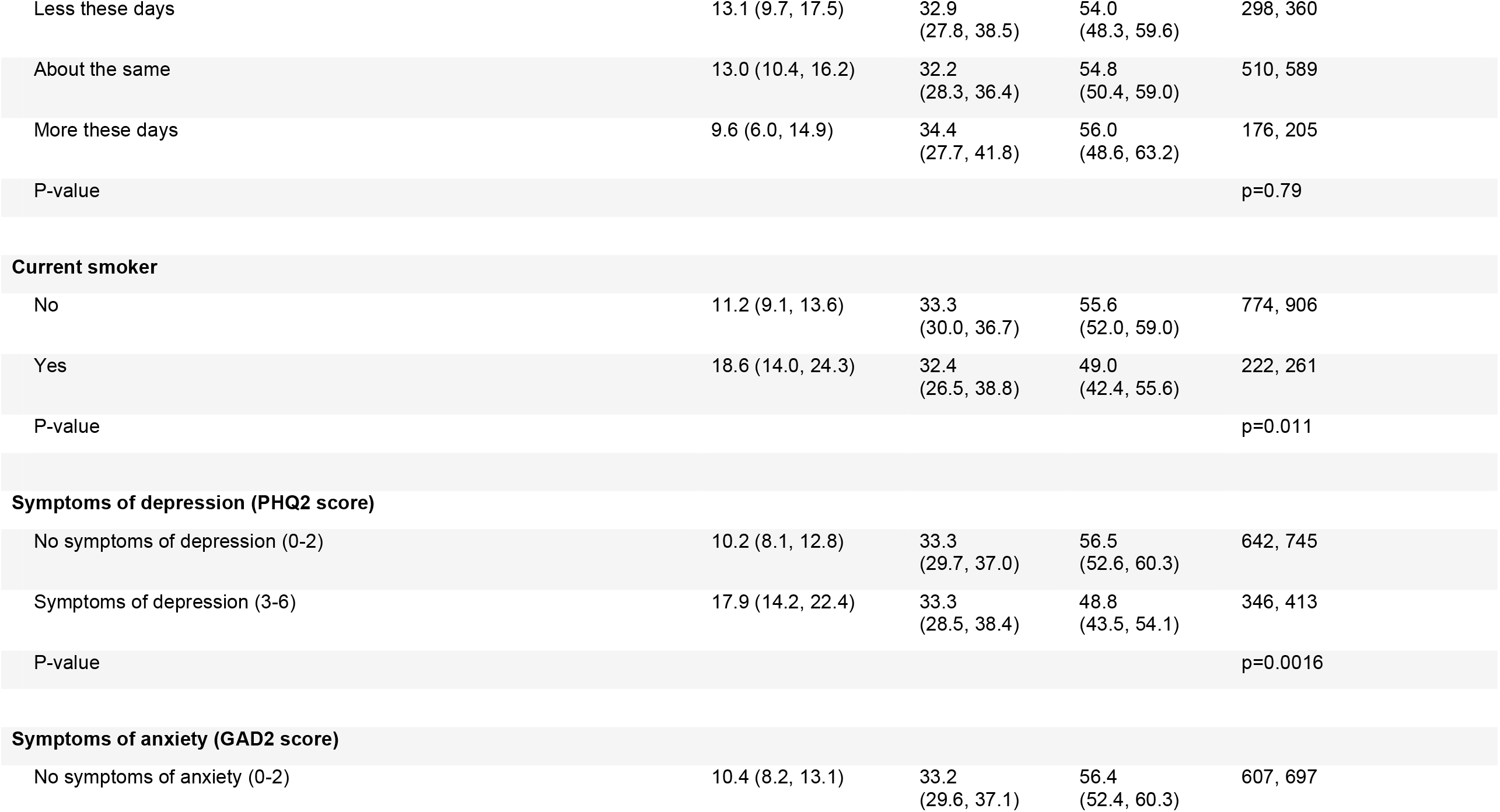
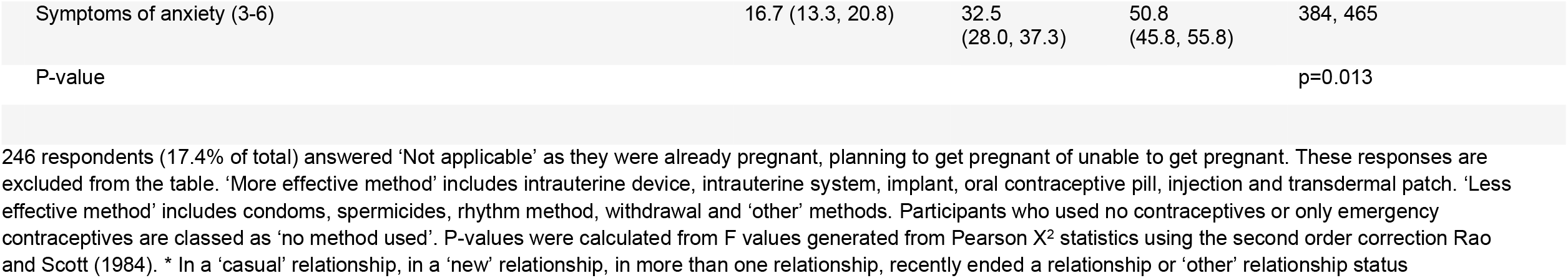
Contraception used in the year since the start of the first UK lockdown by participants aged 18-44 years who were sexually active and were not pregnant, not trying to get pregnant, nor unable to get pregnant.

| Usual contraception used during Covid (% (95%CI)) |  |  |  |  |
| --- | --- | --- | --- | --- |
|  | No method used | Less effective method | More effective method | Denominators (weighted, unweighted) |
| **Total** | 12.8 (10.9, 15.0) | 33.1<br>(30.3, 36.1) | 54.1<br>(51.0, 57.1) | 999, 1169 |
| **Age** |  |  |  |  |
| 18-24 | 12.2 (8.2, 17.8) | 42.0<br>(35.1, 49.3) | 45.7<br>(38.7, 53.0) | 185, 230 |
| 25-29 | 14.9 (11.0, 19.8) | 28.7<br>(23.4, 34.6) | 56.5<br>(50.3, 62.4) | 255, 315 |
| 30-34 | 12.4 (8.6, 17.6) | 33.1<br>(27.1, 39.8) | 54.5<br>(47.7, 61.1) | 212, 252 |

Usual contraception used during Covid (% (95%CI))
|  | No method used | Less effective method | More effective method | Denominators (weighted, unweighted) |
| --- | --- | --- | --- | --- |
| 35-44 | 11.8 (8.8, 15.7) | 31.7<br>(27.0, 36.8) | 56.5<br>(51.2, 61.6) | 346, 372 |
| P-value |  |  |  | p=0.10 |
| Ethnicity |  |  |  |  |
| White | 11.9 (9.9, 14.3) | 30.0<br>(27.0, 33.1) | 58.1<br>(54.7, 61.3) | 863, 1040 |
| Black or Black African or Black Caribbean or Black British | 30.2 (17.0, 47.6) | 42.2<br>(26.8, 59.3) | 27.6<br>(15.1, 45.0) | 35, 27 |
| Asian or Asian British | 19.0 (10.5, 32.0) | 55.1<br>(41.5, 68.0) | 25.9<br>(15.8, 39.4) | 54, 49 |
| Mixed or multiple or other ethnic groups | 10.7 (3.9, 26.2) | 61.8<br>(44.7, 76.4) | 27.5<br>(15.2, 44.7) | 36, 47 |
| P-value |  |  |  | p<0.0001 |
| Self-described sexual identity |  |  |  |  |
| Heterosexual or Straight | 13.0 (11.0, 15.2) | 33.3<br>(30.4, 36.3) | 53.8<br>(50.6, 56.9) | 952, 1035 |
| Lesbian, Gay, Bisexual or Other | 10.2 (3.5, 26.6) | 29.1<br>(16.0, 47.1) | 60.6<br>(42.9, 76.0) | 34, 123 |
| P-value |  |  |  | p=0.40 |
| Social Grade |  |  |  |  |
| A/B - Higher/intermediate managerial, administrative and professional | 11.7 (8.1, 16.5) | 38.1<br>(32.1, 44.6) | 50.2<br>(43.8, 56.6) | 233, 292 |

Usual contraception used during Covid (% (95%CI))
|  | No method used | Less effective method | More effective method | Denominators (weighted, unweighted) |
| --- | --- | --- | --- | --- |
| C1/C2 - Supervisory, clerical and junior managerial, administrative and professional, skilled manual workers | 10.4 (8.1, 13.3) | 28.7 (25.0, 32.8) | 60.9 (56.6, 65.0) | 526, 582 |
| D/E - Semi-skilled and unskilled manual, casual, lowest grade and unemployed | 19.2 (14.7, 24.7) | 37.9 (32.0, 44.3) | 42.9 (36.7, 49.2) | 240, 295 |
| P-value |  |  |  | p<0.0001 |
| Degree | 9.0 (6.8, 11.8) | 36.2 (32.2, 40.5) | 54.8 (50.4, 59.0) | 519, 609 |
| Below degree | 15.3 (12.3, 19.0) | 30.1 (26.0, 34.6) | 54.5 (49.9, 59.1) | 445, 520 |
| No qualifications | 37.4 (22.7, 54.9) | 25.3 (13.3, 42.7) | 37.4 (22.6, 54.9) | 35, 40 |
| P-value |  |  |  | p<0.0001 |
| Living together – relationship but not living together – Single |  |  |  |  |
| Married/steady and living together | 13.5 (11.2, 16.3) | 32.2 (28.8, 35.8) | 54.3 (50.5, 58.0) | 678, 781 |
| Married/steady NOT living together | 10.7 (6.4, 17.3) | 33.6 (25.9, 42.2) | 55.8 (47.1, 64.1) | 131, 164 |
| Casual, new, >1, at end or other | 10.0 (5.0, 19.0) | 42.3 (31.8, 53.5) | 47.7 (36.8, 58.7) | 80, 92 |
| Single | 13.0 (7.8, 20.8) | 31.3 (23.2, 40.7) | 55.7 (46.2, 64.8) | 109, 131 |
| P-value |  |  |  | p=0.64 |

Usual contraception used during Covid (% (95%CI))
|  | No method used | Less effective method | More effective method | Denominators (weighted, unweighted) |
| --- | --- | --- | --- | --- |
| **Been furloughed under the Coronavirus Job Retention Scheme** |  |  |  |  |
| No | 13.0 (10.9, 15.5) | 33.3 (30.2, 36.6) | 53.7 (50.2, 57.1) | 822, 962 |
| Yes | 11.8 (7.8, 17.7) | 33.0 (26.3, 40.5) | 55.2 (47.6, 62.5) | 169, 200 |
| P-value |  |  |  | p=0.90 |
| Became unemployed |  |  |  |  |
| No | 12.8 (10.8, 15.2) | 32.4 (29.4, 35.6) | 54.8 (51.5, 58.0) | 891, 1040 |
| Yes | 12.8 (7.5, 20.9) | 40.8 (31.6, 50.8) | 46.4 (36.8, 56.3) | 101, 122 |
| P-value |  |  |  | p=0.20 |
| Number of days drinking |  |  |  |  |
| 0 days | 15.1 (11.9, 18.9) | 31.6 (27.3, 36.3) | 53.3 (48.4, 58.1) | 409, 474 |
| 1-2 days | 10.6 (7.9, 14.0) | 32.4 (28.0, 37.2) | 57.0 (52.1, 61.8) | 396, 468 |
| 3-4 days | 10.1 (6.0, 16.7) | 41.0 (32.8, 49.7) | 48.8 (40.3, 57.5) | 130, 149 |
| 5-7 days | 16.2 (8.8, 27.8) | 32.3 (21.7, 45.1) | 51.5 (39.0, 63.8) | 62, 76 |
| P-value |  |  |  | p=0.18 |

Drinking habits compared to pre Covid-19 outbreak Usual contraception used during Covid (% (95%CI))
|  | No method used | Less effective method | More effective method | Denominators (weighted, unweighted) |
| --- | --- | --- | --- | --- |
| Less these days | 13.1 (9.7, 17.5) | 32.9 (27.8, 38.5) | 54.0 (48.3, 59.6) | 298, 360 |
| About the same | 13.0 (10.4, 16.2) | 32.2 (28.3, 36.4) | 54.8 (50.4, 59.0) | 510, 589 |
| More these days | 9.6 (6.0, 14.9) | 34.4 (27.7, 41.8) | 56.0 (48.6, 63.2) | 176, 205 |
| P-value |  |  |  | p=0.79 |
| No | 11.2 (9.1, 13.6) | 33.3 (30.0, 36.7) | 55.6 (52.0, 59.0) | 774, 906 |
| Yes | 18.6 (14.0, 24.3) | 32.4 (26.5, 38.8) | 49.0 (42.4, 55.6) | 222, 261 |
| P-value |  |  |  | p=0.011 |
| No symptoms of depression (0-2) | 10.2 (8.1, 12.8) | 33.3 (29.7, 37.0) | 56.5 (52.6, 60.3) | 642, 745 |
| Symptoms of depression (3-6) | 17.9 (14.2, 22.4) | 33.3 (28.5, 38.4) | 48.8 (43.5, 54.1) | 346, 413 |
| P-value |  |  |  | p=0.0016 |
| No symptoms of anxiety (0-2) | 10.4 (8.2, 13.1) | 33.2 (29.6, 37.1) | 56.4 (52.4, 60.3) | 607, 697 |

Usual contraception used during Covid (% (95%CI))
|  | No method used | Less effective method | More effective method | Denominators (weighted, unweighted) |
| --- | --- | --- | --- | --- |
| Symptoms of anxiety (3-6) | 16.7 (13.3, 20.8) | 32.5 (28.0, 37.3) | 50.8 (45.8, 55.8) | 384, 465 |
| P-value |  |  |  | p=0.013 |

246 respondents (17.4% of total) answered 'Not applicable' as they were already pregnant, planning to get pregnant or unable to get pregnant. These responses are excluded from the table. 'More effective method' includes intrauterine device, intrauterine system, implant, oral contraceptive pill, injection and transdermal patch. 'Less effective method' includes condoms, spermicides, rhythm method, withdrawal and 'other' methods. Participants who used no contraceptives or only emergency contraceptives are classed as 'no method used'. P-values were calculated from F values generated from Pearson $\chi^2$ statistics using the second order correction Rao and Scott (1984). \* In a 'casual' relationship, in a 'new' relationship, in more than one relationship, recently ended a relationship or 'other' relationship status

Overall, 12.8% (10.9%-15.0%) of participants at risk of an unplanned pregnancy reported no usual contraception methods (Table 2). This was more likely in those who reported smoking (p=0.011), lower educational qualification (p<0.0001) and poor mental health (depression: p=0.0016; anxiety: p=0.013).

In total, 227 participants (14.3% (12.5%-16.3%) of participants included in this analysis) reported stopping or switching contraceptive method due to the pandemic. Among those using a more effective contraceptive at the start of the pandemic, 10.2% (7.9%-13.1%) reported switching to a similarly or more effective method, 3.2% (2.0%-5.1%) switched to a less effective method and 3.8% (2.5%-5.9%) stopped (Table 3). Among users of effective methods, we found differences in stopping/switching by age, ethnicity and sociodemographic factors. Those aged 18–24 were more likely than older participants to have switched method (23.8% (15.4%-34.8%) versus aged 25–44: 11.7% (9.0%-15.0%)). Compared with white participants, Black participants were more likely to have switched their usual method (29.6% (9.2%-63.7%) versus 11.8% (9.4%-14.8%)) and to have stopped using contraceptives (9.7% (1.2%-49.3%) versus 3.7% (2.3%-5.8%)). Reporting depression was associated with switching method (20.0% (14.6%-26.8%) versus 10.0% (7.2%-13.6%); Table 3).

**Table 3.**
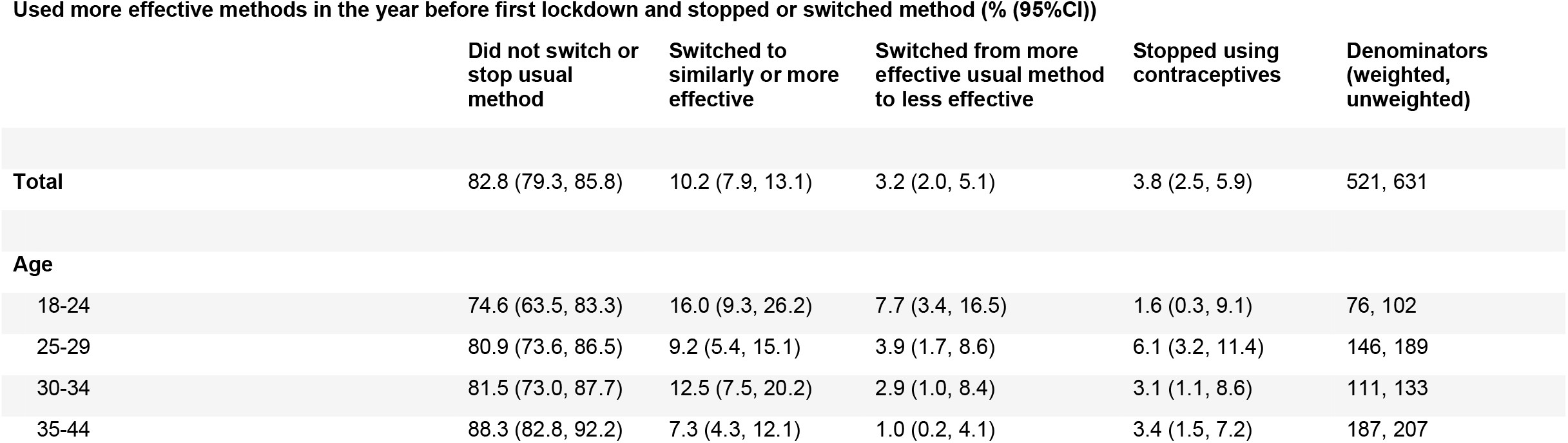
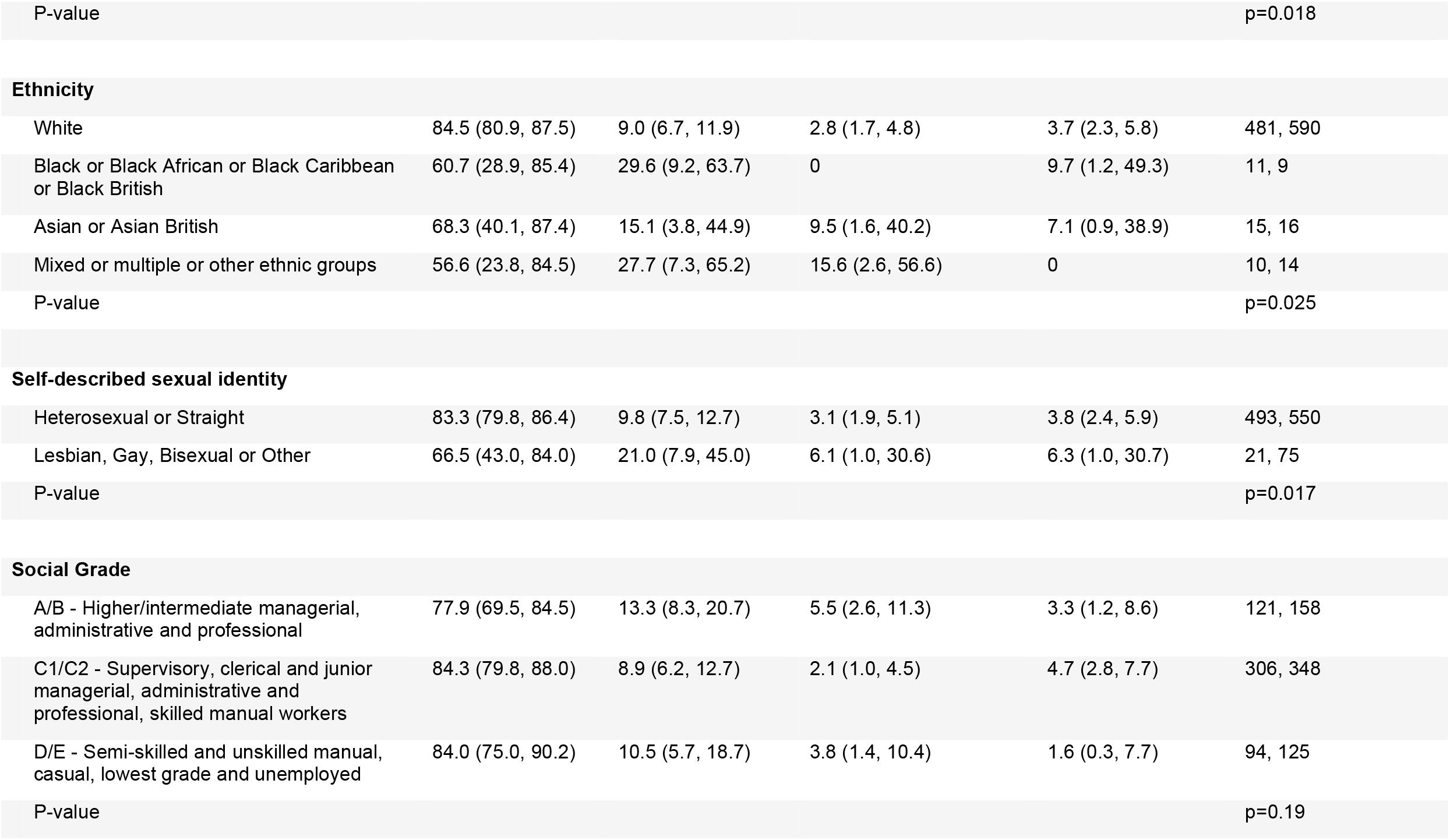
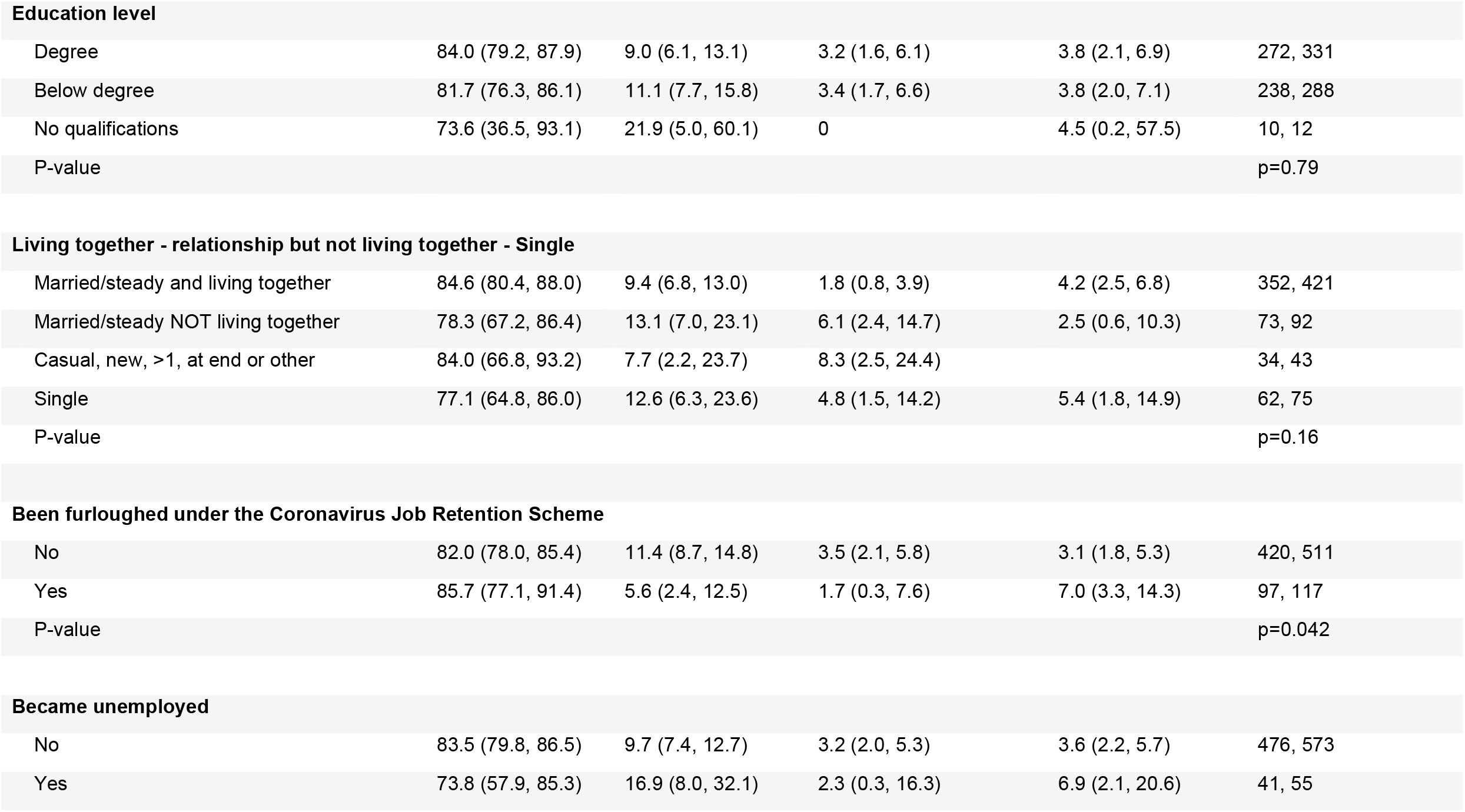
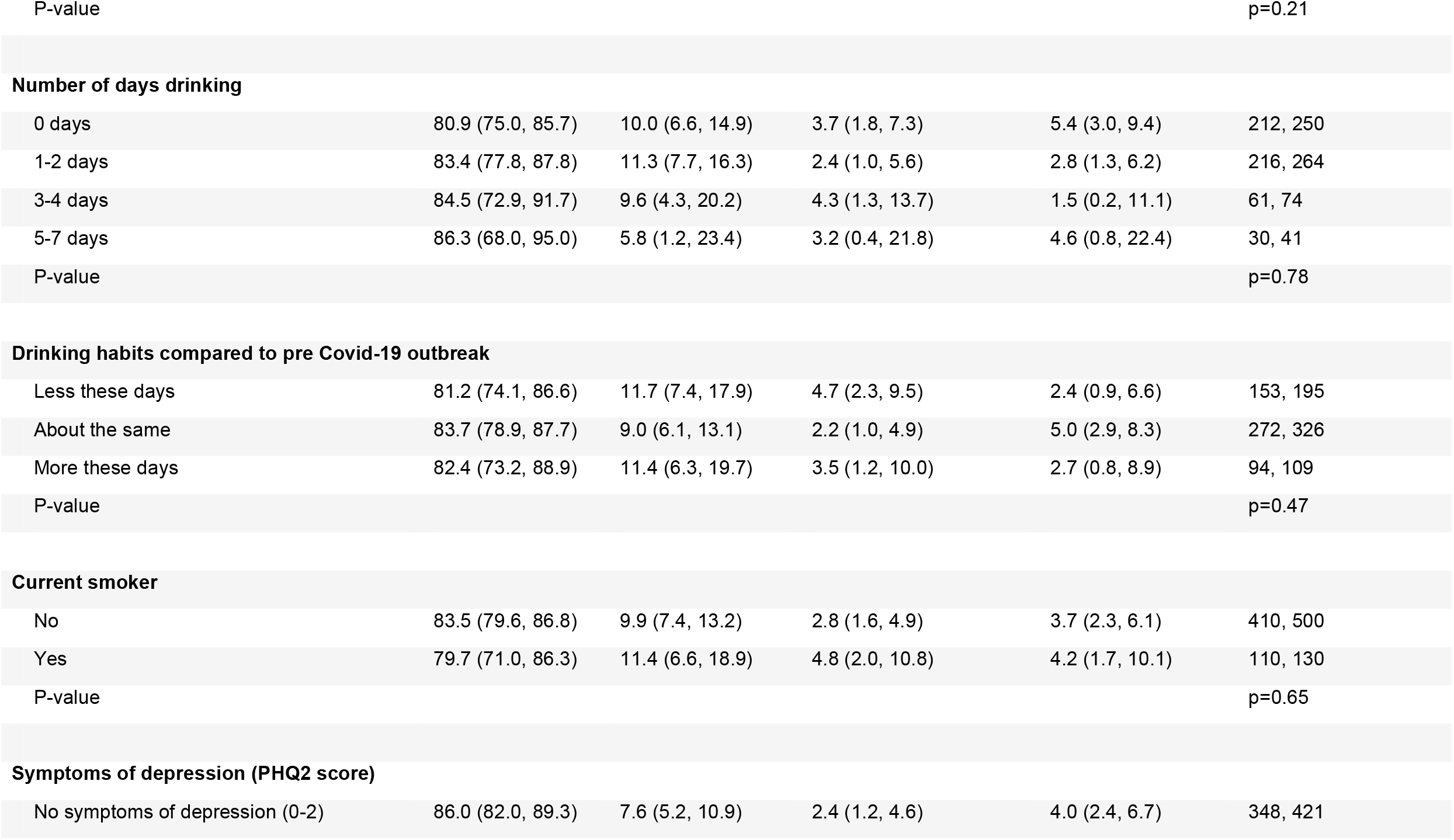
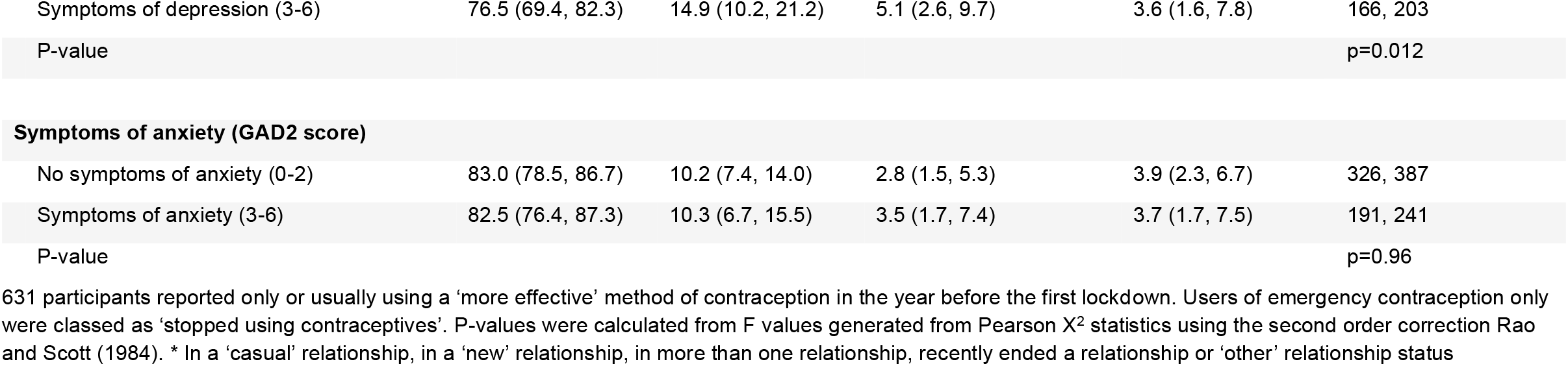
Switching due to the pandemic from usual pre-COVID contraception method, among participants who were using ‘more effective’ contraceptives and were not pregnant, not trying to get pregnant, nor unable to get pregnant.

Used more effective methods in the year before first lockdown and stopped or switched method (% (95%CI))
|  | Did not switch or stop usual method | Switched to similarly or more effective | Switched from more effective usual method to less effective | Stopped using contraceptives | Denominators (weighted, unweighted) |
| --- | --- | --- | --- | --- | --- |
| **Total** | 82.8 (79.3, 85.8) | 10.2 (7.9, 13.1) | 3.2 (2.0, 5.1) | 3.8 (2.5, 5.9) | 521, 631 |
| **Age** |  |  |  |  |  |
| 18-24 | 74.6 (63.5, 83.3) | 16.0 (9.3, 26.2) | 7.7 (3.4, 16.5) | 1.6 (0.3, 9.1) | 76, 102 |
| 25-29 | 80.9 (73.6, 86.5) | 9.2 (5.4, 15.1) | 3.9 (1.7, 8.6) | 6.1 (3.2, 11.4) | 146, 189 |
| 30-34 | 81.5 (73.0, 87.7) | 12.5 (7.5, 20.2) | 2.9 (1.0, 8.4) | 3.1 (1.1, 8.6) | 111, 133 |
| 35-44 | 88.3 (82.8, 92.2) | 7.3 (4.3, 12.1) | 1.0 (0.2, 4.1) | 3.4 (1.5, 7.2) | 187, 207 |

Used more effective methods in the year before first lockdown and stopped or switched method (% (95%CI))
|  | Did not switch or stop usual method | Switched to similarly or more effective | Switched from more effective usual method to less effective | Stopped using contraceptives | Denominators (weighted, unweighted) |
| --- | --- | --- | --- | --- | --- |
| P-value |  |  |  |  | p=0.018 |
| **Ethnicity** |  |  |  |  |  |
| White | 84.5 (80.9, 87.5) | 9.0 (6.7, 11.9) | 2.8 (1.7, 4.8) | 3.7 (2.3, 5.8) | 481, 590 |
| Black or Black African or Black Caribbean or Black British | 60.7 (28.9, 85.4) | 29.6 (9.2, 63.7) | 0 | 9.7 (1.2, 49.3) | 11, 9 |
| Asian or Asian British | 68.3 (40.1, 87.4) | 15.1 (3.8, 44.9) | 9.5 (1.6, 40.2) | 7.1 (0.9, 38.9) | 15, 16 |
| Mixed or multiple or other ethnic groups | 56.6 (23.8, 84.5) | 27.7 (7.3, 65.2) | 15.6 (2.6, 56.6) | 0 | 10, 14 |
| P-value |  |  |  |  | p=0.025 |
| **Self-described sexual identity** |  |  |  |  |  |
| Heterosexual or Straight | 83.3 (79.8, 86.4) | 9.8 (7.5, 12.7) | 3.1 (1.9, 5.1) | 3.8 (2.4, 5.9) | 493, 550 |
| Lesbian, Gay, Bisexual or Other | 66.5 (43.0, 84.0) | 21.0 (7.9, 45.0) | 6.1 (1.0, 30.6) | 6.3 (1.0, 30.7) | 21, 75 |
| P-value |  |  |  |  | p=0.017 |
| **Social Grade** |  |  |  |  |  |
| A/B - Higher/intermediate managerial, administrative and professional | 77.9 (69.5, 84.5) | 13.3 (8.3, 20.7) | 5.5 (2.6, 11.3) | 3.3 (1.2, 8.6) | 121, 158 |
| C1/C2 - Supervisory, clerical and junior managerial, administrative and professional, skilled manual workers | 84.3 (79.8, 88.0) | 8.9 (6.2, 12.7) | 2.1 (1.0, 4.5) | 4.7 (2.8, 7.7) | 306, 348 |
| D/E - Semi-skilled and unskilled manual, casual, lowest grade and unemployed | 84.0 (75.0, 90.2) | 10.5 (5.7, 18.7) | 3.8 (1.4, 10.4) | 1.6 (0.3, 7.7) | 94, 125 |
| P-value |  |  |  |  | p=0.19 |

Used more effective methods in the year before first lockdown and stopped or switched method (% (95%CI))
|  | Did not switch or stop usual method | Switched to similarly or more effective | Switched from more effective usual method to less effective | Stopped using contraceptives | Denominators (weighted, unweighted) |
| --- | --- | --- | --- | --- | --- |
| **Education level** |  |  |  |  |  |
| Degree | 84.0 (79.2, 87.9) | 9.0 (6.1, 13.1) | 3.2 (1.6, 6.1) | 3.8 (2.1, 6.9) | 272, 331 |
| Below degree | 81.7 (76.3, 86.1) | 11.1 (7.7, 15.8) | 3.4 (1.7, 6.6) | 3.8 (2.0, 7.1) | 238, 288 |
| No qualifications | 73.6 (36.5, 93.1) | 21.9 (5.0, 60.1) | 0 | 4.5 (0.2, 57.5) | 10, 12 |
| P-value |  |  |  |  | p=0.79 |
| **Living together - relationship but not living together - Single** |  |  |  |  |  |
| Married/steady and living together | 84.6 (80.4, 88.0) | 9.4 (6.8, 13.0) | 1.8 (0.8, 3.9) | 4.2 (2.5, 6.8) | 352, 421 |
| Married/steady NOT living together | 78.3 (67.2, 86.4) | 13.1 (7.0, 23.1) | 6.1 (2.4, 14.7) | 2.5 (0.6, 10.3) | 73, 92 |
| Casual, new, >1, at end or other | 84.0 (66.8, 93.2) | 7.7 (2.2, 23.7) | 8.3 (2.5, 24.4) |  | 34, 43 |
| Single | 77.1 (64.8, 86.0) | 12.6 (6.3, 23.6) | 4.8 (1.5, 14.2) | 5.4 (1.8, 14.9) | 62, 75 |
| P-value |  |  |  |  | p=0.16 |
| **Been furloughed under the Coronavirus Job Retention Scheme** |  |  |  |  |  |
| No | 82.0 (78.0, 85.4) | 11.4 (8.7, 14.8) | 3.5 (2.1, 5.8) | 3.1 (1.8, 5.3) | 420, 511 |
| Yes | 85.7 (77.1, 91.4) | 5.6 (2.4, 12.5) | 1.7 (0.3, 7.6) | 7.0 (3.3, 14.3) | 97, 117 |
| P-value |  |  |  |  | p=0.042 |
| **Became unemployed** |  |  |  |  |  |
| No | 83.5 (79.8, 86.5) | 9.7 (7.4, 12.7) | 3.2 (2.0, 5.3) | 3.6 (2.2, 5.7) | 476, 573 |
| Yes | 73.8 (57.9, 85.3) | 16.9 (8.0, 32.1) | 2.3 (0.3, 16.3) | 6.9 (2.1, 20.6) | 41, 55 |

Used more effective methods in the year before first lockdown and stopped or switched method (% (95%CI))
|  | Did not switch or stop usual method | Switched to similarly or more effective | Switched from more effective usual method to less effective | Stopped using contraceptives | Denominators (weighted, unweighted) |
| --- | --- | --- | --- | --- | --- |
| P-value |  |  |  |  | p=0.21 |
| Number of days drinking |  |  |  |  |  |
| 0 days | 80.9 (75.0, 85.7) | 10.0 (6.6, 14.9) | 3.7 (1.8, 7.3) | 5.4 (3.0, 9.4) | 212, 250 |
| 1-2 days | 83.4 (77.8, 87.8) | 11.3 (7.7, 16.3) | 2.4 (1.0, 5.6) | 2.8 (1.3, 6.2) | 216, 264 |
| 3-4 days | 84.5 (72.9, 91.7) | 9.6 (4.3, 20.2) | 4.3 (1.3, 13.7) | 1.5 (0.2, 11.1) | 61, 74 |
| 5-7 days | 86.3 (68.0, 95.0) | 5.8 (1.2, 23.4) | 3.2 (0.4, 21.8) | 4.6 (0.8, 22.4) | 30, 41 |
| P-value |  |  |  |  | p=0.78 |
| Less these days | 81.2 (74.1, 86.6) | 11.7 (7.4, 17.9) | 4.7 (2.3, 9.5) | 2.4 (0.9, 6.6) | 153, 195 |
| About the same | 83.7 (78.9, 87.7) | 9.0 (6.1, 13.1) | 2.2 (1.0, 4.9) | 5.0 (2.9, 8.3) | 272, 326 |
| More these days | 82.4 (73.2, 88.9) | 11.4 (6.3, 19.7) | 3.5 (1.2, 10.0) | 2.7 (0.8, 8.9) | 94, 109 |
| P-value |  |  |  |  | p=0.47 |
| No | 83.5 (79.6, 86.8) | 9.9 (7.4, 13.2) | 2.8 (1.6, 4.9) | 3.7 (2.3, 6.1) | 410, 500 |
| Yes | 79.7 (71.0, 86.3) | 11.4 (6.6, 18.9) | 4.8 (2.0, 10.8) | 4.2 (1.7, 10.1) | 110, 130 |
| P-value |  |  |  |  | p=0.65 |
| No symptoms of depression (0-2) | 86.0 (82.0, 89.3) | 7.6 (5.2, 10.9) | 2.4 (1.2, 4.6) | 4.0 (2.4, 6.7) | 348, 421 |

**Used more effective methods in the year before first lockdown and stopped or switched method (% (95%CI))**
|  | **Did not switch or stop usual method** | **Switched to similarly or more effective** | **Switched from more effective usual method to less effective** | **Stopped using contraceptives** | **Denominators (weighted, unweighted)** |
| --- | --- | --- | --- | --- | --- |
| Symptoms of depression (3-6) | 76.5 (69.4, 82.3) | 14.9 (10.2, 21.2) | 5.1 (2.6, 9.7) | 3.6 (1.6, 7.8) | 166, 203 |
| P-value |  |  |  |  | p=0.012 |
| **Symptoms of anxiety (GAD2 score)** |  |  |  |  |  |
| No symptoms of anxiety (0-2) | 83.0 (78.5, 86.7) | 10.2 (7.4, 14.0) | 2.8 (1.5, 5.3) | 3.9 (2.3, 6.7) | 326, 387 |
| Symptoms of anxiety (3-6) | 82.5 (76.4, 87.3) | 10.3 (6.7, 15.5) | 3.5 (1.7, 7.4) | 3.7 (1.7, 7.5) | 191, 241 |
| P-value |  |  |  |  | p=0.96 |

631 participants reported only or usually using a 'more effective' method of contraception in the year before the first lockdown. Users of emergency contraception only were classed as 'stopped using contraceptives'. P-values were calculated from F values generated from Pearson X<sup>2</sup> statistics using the second order correction Rao and Scott (1984). \* In a 'casual' relationship, in a 'new' relationship, in more than one relationship, recently ended a relationship or 'other' relationship status

Unmet needs of contraceptive services varied by sexual identity and markers of physical and mental health. Amongst all participants (n=1,488), 29.3% (26.9%-31.8%) reported trying to access a contraceptive service between March 2020 and April 2021; 74 (16.4% (13.0%-20.4%) of those who tried to access) reported being unable to do so at least once (unmet need). Many of those also reported at least one successful access attempt; only 24 were unable to access a contraceptive service at all (Figure 1a; Supplementary Table 2). Young participants were most likely to report an unmet need (7.4% (6.1%-9.0%) compared with those aged 35–44 years: 2.9% (2.1%-3.9%); Figure 2), as were those from minoritised sexual identities (11.7% (10.0%-13.6%) compared with heterosexuals: 4.5% (3.5%-5.8%)).

**Figure 1.png.**
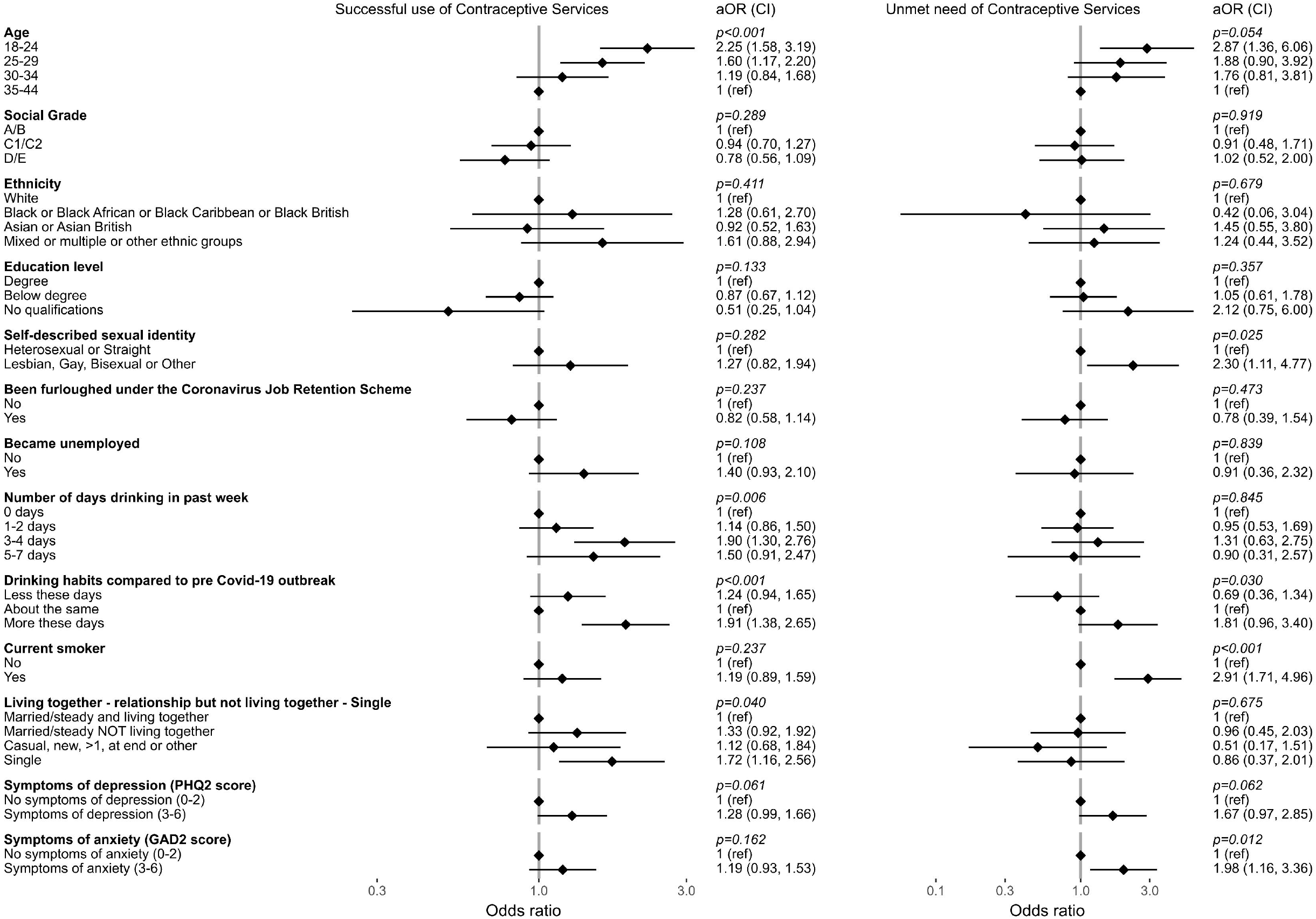
Accessing contraception services

**Figure 2.png.**
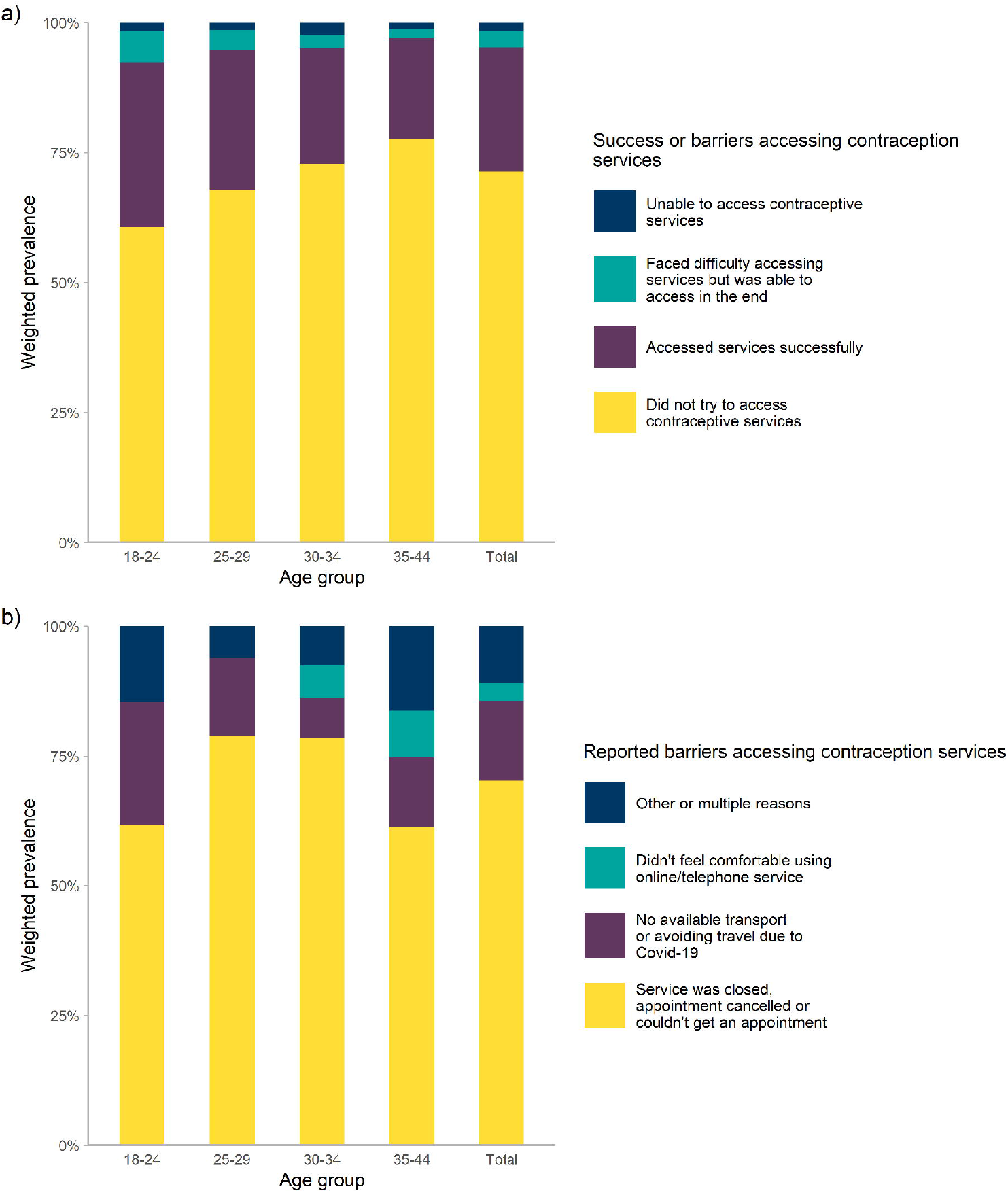
Contraception service access outcomes amongst all participants (n=1,488) – factors associated with success and unmet need Footnote: All ORs are age-adjusted, except those for the age categories which are crude. Analyses are conducted across 441 participants (29.6%) who attempted to access a contraceptive service at least once since the start of the first lockdown. Social grade codes: A/B - Higher/intermediate managerial, administrative and professional; C1/C2 - Supervisory, clerical and junior managerial, administrative and professional, skilled manual workers; D/E - Semi-skilled and unskilled manual, casual, lowest grade and unemployed

After adjustment for age, anxiety (aOR: 1.98 (1.16-3.36)) and depression (aOR: 1.67 (0.97-2.85)) were both associated with unmet need. Current smokers were also at higher risks of unmet need (aOR: 2.91 (1.71-4.96) vs non-smokers). Of barriers cited by those with unmet need (n= 74), most related to clinic closures and appointment cancellations or unavailability (70.2%, Figure 1b).

Among all participants (n=1,488), 13.6% reported a current pregnancy or pregnancy in the past year (n=199). The mean LMUP score for these pregnancies was 9.2 (SD: 3.0; with scores >9 classed as ‘planned’), and 6.6% (3.9%-11.1%) were scored as an unplanned pregnancy (LMUP score, 0–3). By comparison, among the 285 participants who reported a pregnancy between 1 and 5 years ago (but no pregnancy in the past year), the mean LMUP score was 8.6 (SD: 3.6; difference in weighted mean = 0.58, p = 0.064; data not shown), and 12.3% (8.8%-16.8%) were scored as unplanned. Eleven participants (6.1% of those reporting a pregnancy) reported an abortion in the past year, and six of these had a pregnancy that was scored as unplanned, though none of these participants reported unsuccessful attempts to access contraceptive services (Table 4). Pregnancies in older participants were more likely than those in younger participants to be more planned (difference in mean score for those aged 25–29: 2.90 (1.41-4.40), compared with participants aged 18–24, Table 4). Cohabitation, relationship status and social grade were associated with reported pregnancy and pregnancy planning scores. Pregnancies were less commonly reported by those in non-cohabiting relationships compared with those living with their partner (7.3% (4.1%-12.7%) versus 17.1% (14.8%-19.7%) respectively). Single participants were less likely to report being pregnant (3.0% (1.1%-8.0%)), but those who did were more likely to have an unplanned pregnancy compared with those in a steady or married non-cohabiting relationship (age-adjusted difference compared to single participants: 4.46 (2.39-6.53)) or a cohabiting relationship (age-adjusted difference: 5.31 (3.61-7.00)). Those working in less-skilled occupations, receiving state benefit or unemployed had lower LMUP scores. Smoking was associated with lower LMUP scores (age-adjusted score difference: -1.10 (−2.16 to -0.05)).

**Table 4.**
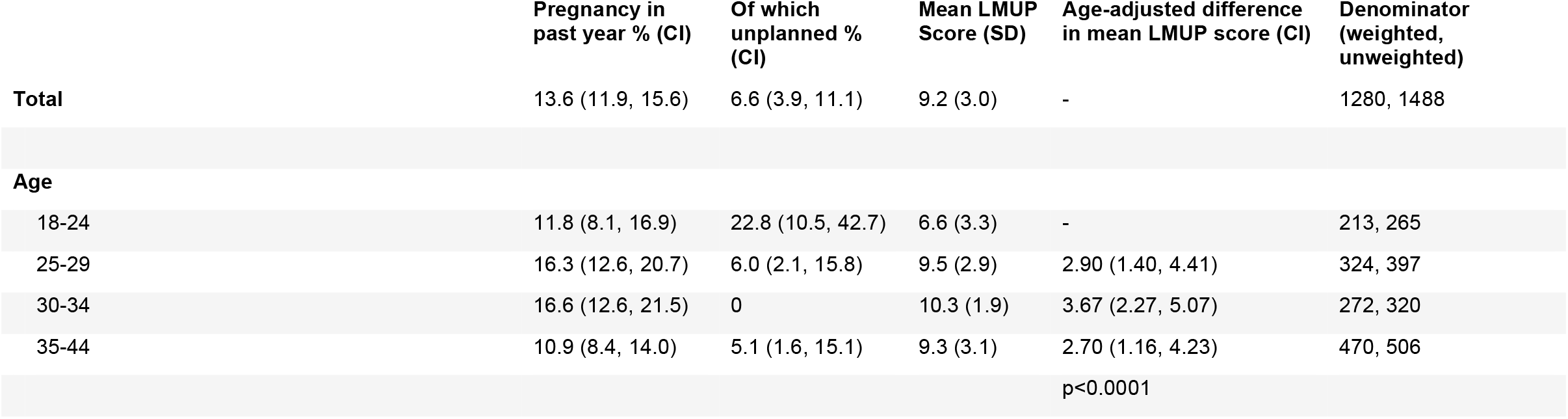
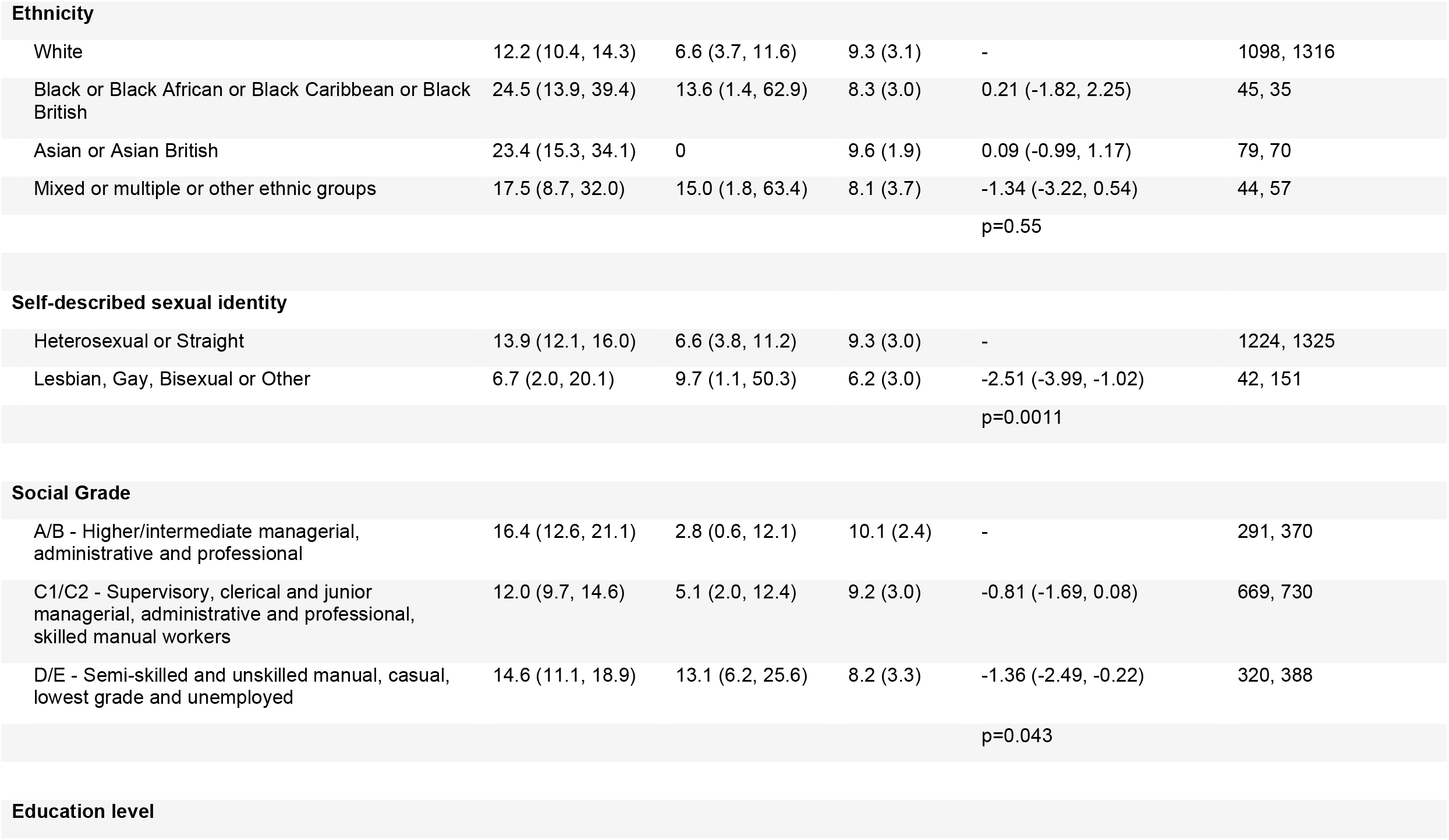
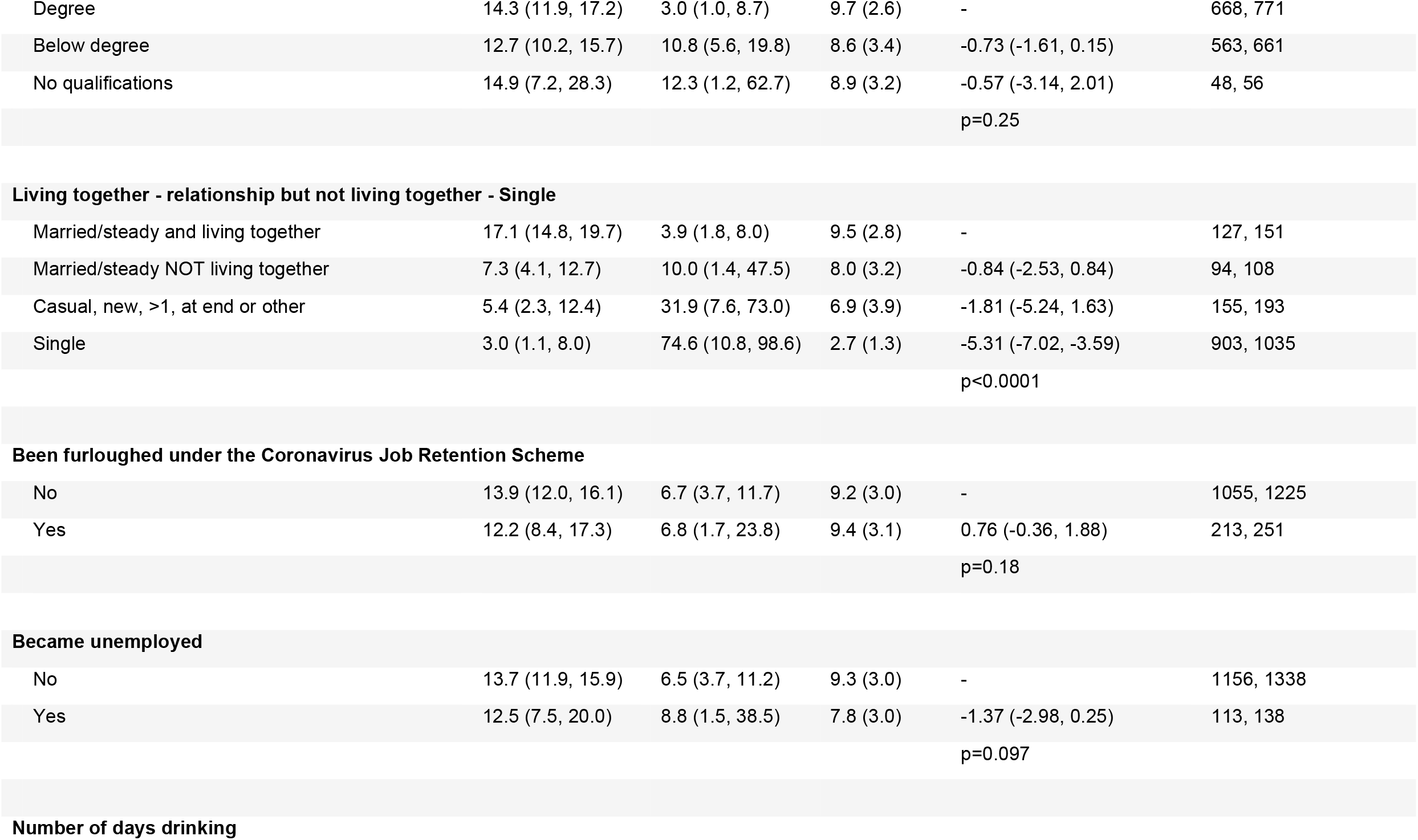
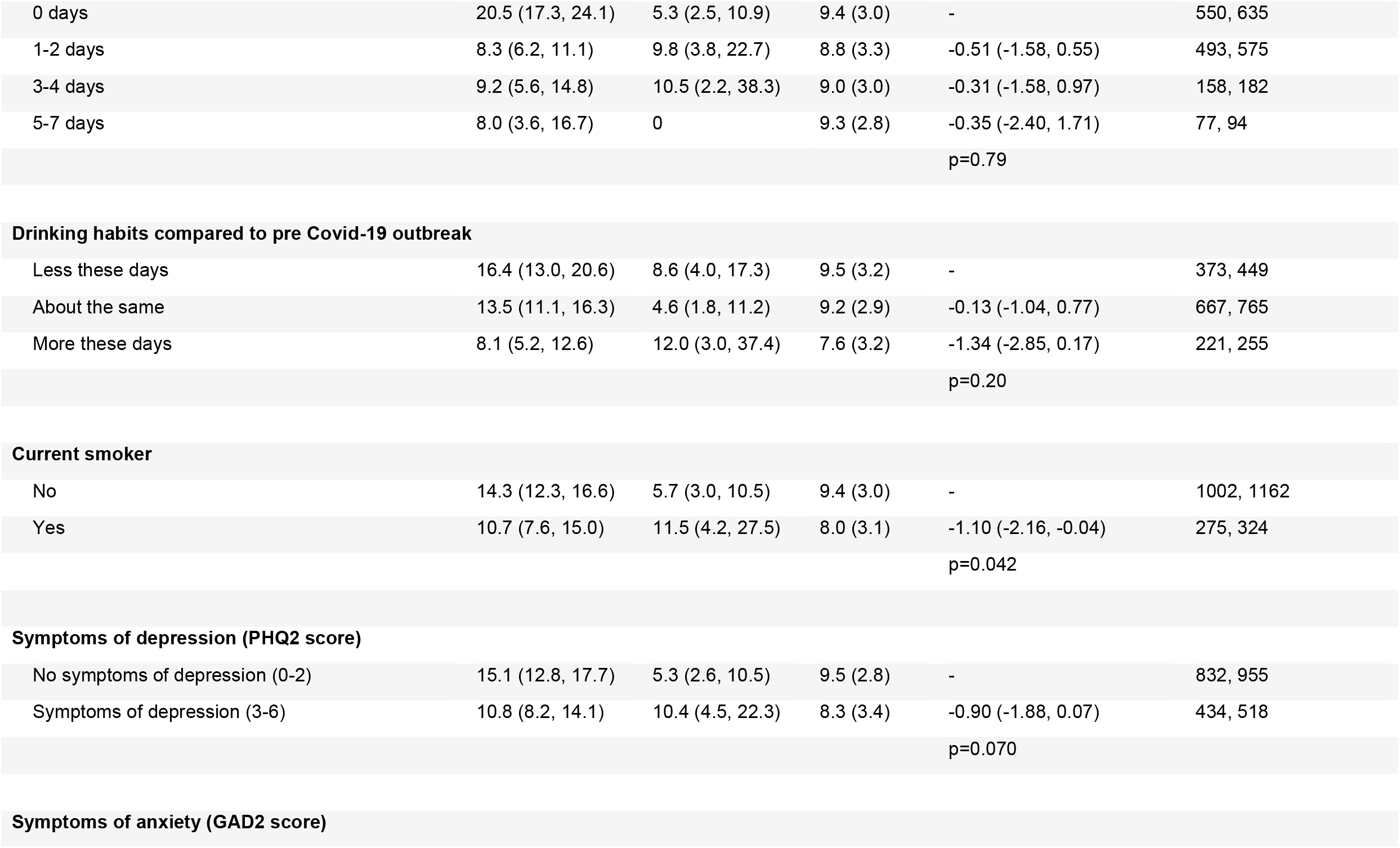
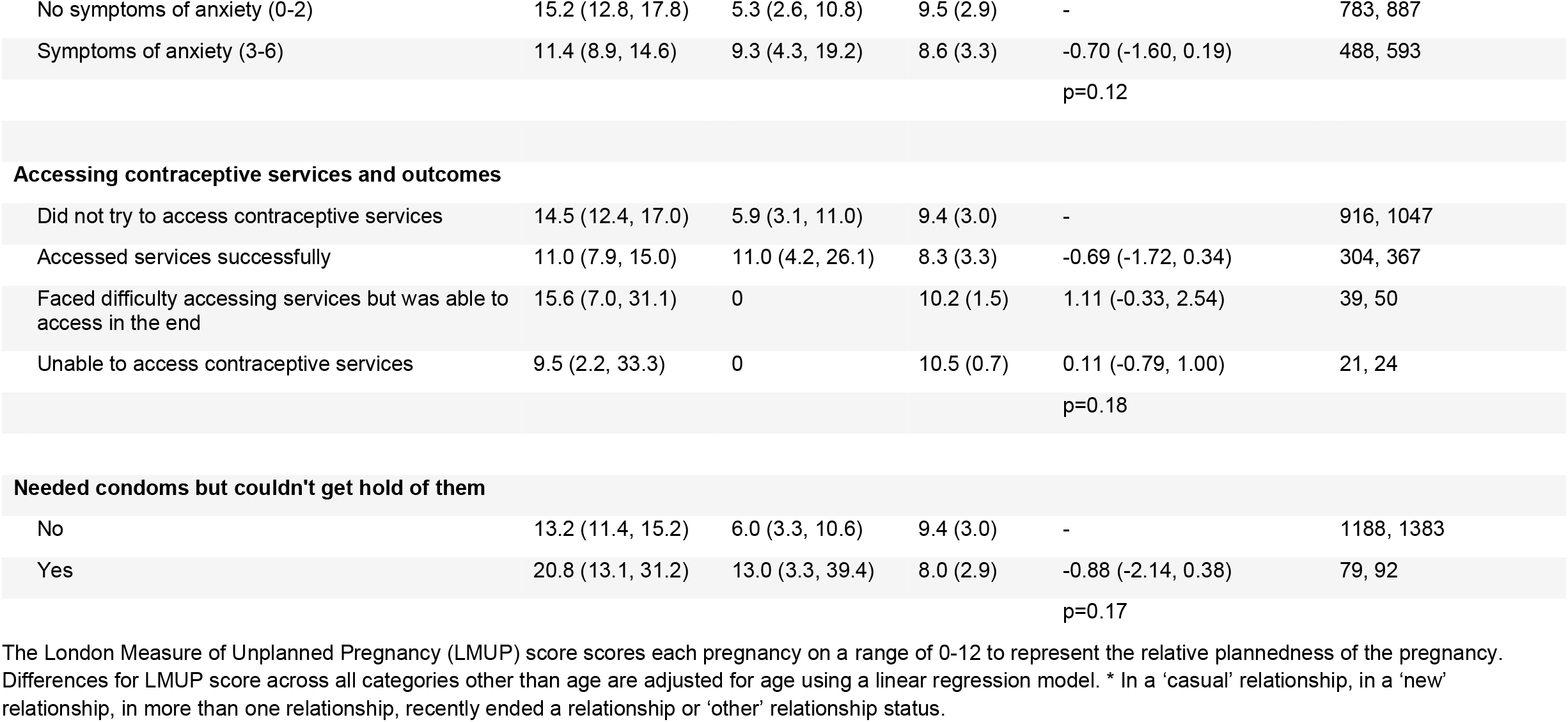
Pregnancies in the past year and their ‘plannedness’ scored using the London measure of unplanned pregnancy (LMUP) among sexually active participants aged 18-44 years.

## Discussion

Our study used a large quasi-representative sample of the British general population and emphasises the high level of need for contraceptive services that continued during the year after the first national COVID-19 lockdown. The finding that most participants reported being able to access the services they needed indicates resilience in service delivery. Moreover, only 16.4% of those attempting to access a contraceptive service reported an unsuccessful attempt and most of these also reported a successful attempt. Overall, 53.9% of participants reported using an effective method as their usual contraception during the pandemic, and this proportion was similar to the 56.4% and 54.2% using effective methods found in previous Natsal surveys: the Natsal-2 (2000–2001) and Natsal-3 (2010–2012) surveys respectively.^31^ However, we found that low proportions of participants from Asian or Asian-British ethnic backgrounds (25.9%) or from a mixed or multiple or other ethnic background (27.5%), compared to participants from White ethnic backgrounds (58.1%) reported using effective methods. Though likely due at least partly to pre-pandemic differences,^32^ this suggests different levels of risk for unplanned pregnancy by ethnicity during this national period of high stress and uncertainty. It was reassuring that most participants (82.8%) using effective contraception pre-pandemic reported not having to switch method or stop using contraception because of the pandemic, and 10.2% reported switching but were able to use similarly or more effective methods. However, consistent with earlier research, we found that younger participants (most at risk of unplanned pregnancy) were more likely to have switched method because of the pandemic, and to report barriers to accessing contraceptive services.^17^ Routinely collected data indicate large reductions in contraception prescription and dispensing in England in 2020 compared to 2019.^33–35^ Our data suggest difficulties accessing services, primarily due to closures and appointment cancellations, may have contributed to this reduction.

Our analysis of pregnancies during the pandemic builds upon and challenges previous research. Elsewhere we report that, compared to Natsal-3 data collected a decade ago (in 2010-12), pregnancies and abortions in the first year of the pandemic were substantially lower.^19^ We also found that pregnancies during the pandemic were less likely to be scored as unplanned compared to a decade previously (6.2% vs. 18.3%)^19^, which is consistent with our Natsal-COVID data on participants’ pregnancies reported in the four years before the pandemic (12.3% scored as unplanned). On the one hand, these observations might be explained by improvements in service provision impacting on access to contraceptive methods, especially long-acting reversible contraceptives. On the other hand, less sexual contact during the pandemic might have led to lower pregnancy rates, ^36^ reducing risk of unplanned pregnancy regardless of contraception and service access. The narrowing of fertility-rate gaps across deprivation quintiles also suggests declines were primarily driven by falling rates of unplanned pregnancy.^37^

In contrast to our findings indicating a lower proportion of unplanned pregnancies during the pandemic, a convenience sample cohort study of pregnant women in the UK found that conceptions in the year following the first lockdown were more likely to be unplanned than pre-lockdown conceptions.^14^ Two study design factors might explain the different results. The cohort study used online adverts to recruit participants, which might introduce bias, and only included participants who were still pregnant at time of recruitment (commencing May 2020), thus excluding unplanned pregnancy terminations before the end of April 2020. The Natsal-COVID estimate, recruiting a wider range of participants and using weighting to achieve representativeness, seems less susceptible to bias.

In our quasi-representative population sample, several markers of vulnerability and health risk behaviours were associated with elevated risks of contraception switching, unmet need of contraceptive services and unplanned pregnancy. Whilst patterns of reproductive health risks during the pandemic may match existing inequalities, pandemic-induced inequalities in access to contraceptive services may have exacerbated these. Participants with poor health and behavioural risk factors such as smoking and drinking alcohol reported higher rates of unmet need for contraceptive services and higher rates of switching or stopping contraceptives. While these findings suggest a greater risk of unplanned pregnancy in these groups, we were unable to directly link the ‘plannedness’ of each pregnancy to specific attempts to access contraceptive services. Instead, it was those in lower social grades and who smoked who were more likely to reported unplanned pregnancies, which is similar to patterns previously observed in the UK.^7,37^ In our study, most people switching contraceptive method switched to a similarly, or more, effective method, suggesting flexibility and adaptability in participants’ responses to changing service provision, which might have been sufficient to meet contraceptive needs in many cases. Our findings are also consistent with convenience-sample evidence from the USA that a drop in desire to achieve pregnancy was associated with low income, but not independently associated with decreased income due to the pandemic.^38^ Natsal-COVID benefited from a questionnaire design and approach developed by the team responsible for the decennial Natsal survey to obtain rigorous data on potentially sensitive behaviours and experiences.^20^ Natsal-COVID included a large, national sample and used quota-based sampling and weighting to improve representativeness. Unlike the decennial Natsal survey, Natsal-COVID was not a probability sample, and is therefore not directly representative of the general population.^39,40^ In several cases, in particular among sexual minority people, and trans and non-binary people, small numbers of participants in sub-categories precluded the testing of distinct inequalities across these groups. Small numbers of participants from some ethnic groups limited our ability to detect differences in outcomes.

Our study informs adaptations to contraceptive services to meet patient needs and preferences, including in the recovery phase from the COVID-19 pandemic. Regardless of differences in how health systems are structured, financed or commissioned in other high-income countries, our findings broadly indicate the likely impacts of the pandemic on contraceptive method and service use. We highlight here inequalities across age, ethnicity, social disadvantage and mental health. Ongoing provision of contraceptive services and future crisis planning should ensure ease and equality of access to contraceptive services for all to address the impact on contraceptive method choice and availability.

## Data Availability

All data produced will be made available online at https://www.natsal.ac.uk/resources/accessing-data (undergoing preparation at time of preprint)

## Boxes

### Box 1 Classifications of contraceptive types

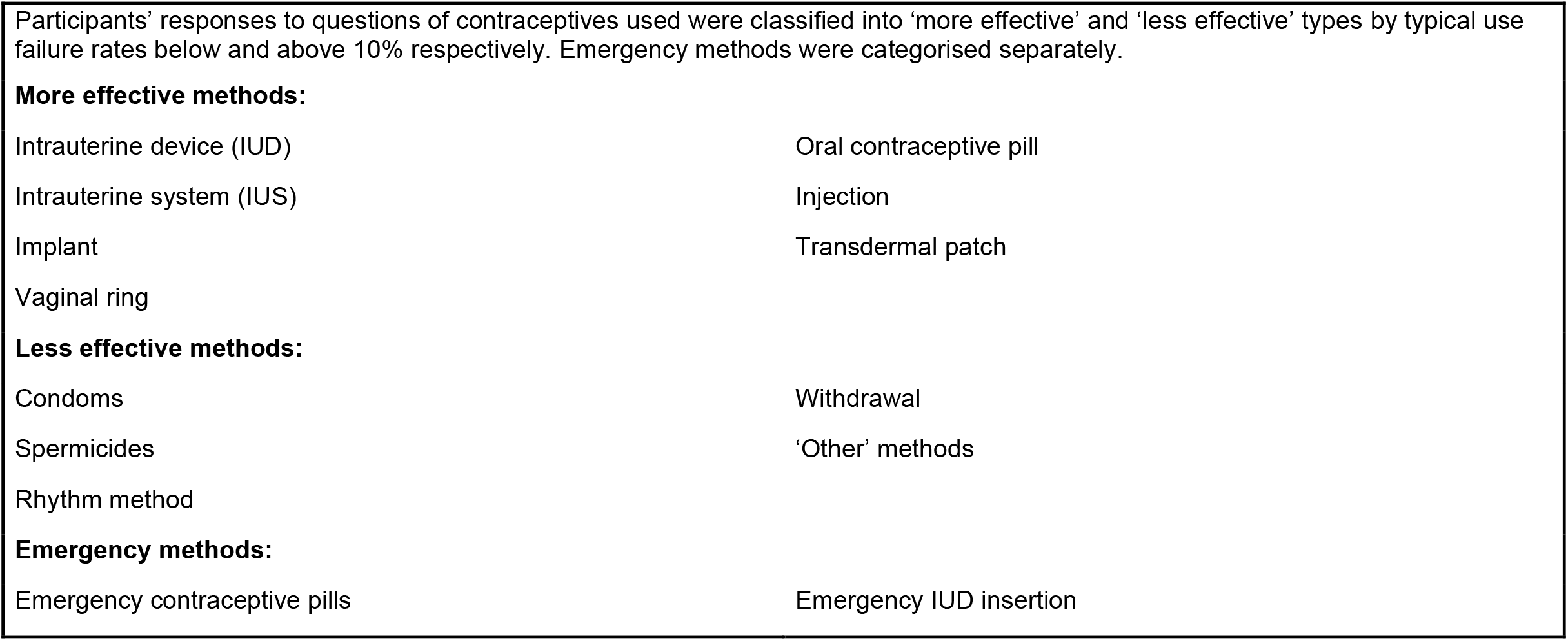

## Notes

The authors declare no conflicts of interest.

### Competing Interest Statement

The authors have declared no competing interest.

### Funding Statement

The study was funded by the Wellcome Trust, The Economic and Social Research Council, The National Institute for Health Research, Medical Research Council/Chief Scientist Office Social and Public Health Sciences Unit, and UCL Coronavirus Response Fund.

### Author Declarations

Ethics committee of University of Glasgow gave ethical approval for this work. Ethics committee of London School of Hygiene and Tropical Medicine gave ethical approval for this work.

