## Supplementary Table for "Contraceptive use and pregnancy planning in Britain during the first year of the COVID-19 pandemic: findings from a large, quasi-representative survey (Natsal-COVID)"

### Supplementary Material

#### Supplementary Table 1: Definitions of outcome variables

| Outcome/Variable | Denominator | Definition |
| --- | --- | --- |
| Usual contraception used in the year before lockdown | At risk of unplanned pregnancy (n=1,169) | Selected contraceptive method in response to “In the year before the start of the first lockdown (23 March 2020), which of the following did you or a partner use to prevent pregnancy?”. Participants selecting more than one method were asked to specify their ‘usual’ method. |
| Usual contraception used in the year since lockdown started | At risk of unplanned pregnancy (n=1,169) | Selected contraceptive method in response to “Since the start of the first lockdown, which of the following did you or a partner use to prevent pregnancy?”. Participants selecting more than one method were asked to specify their ‘usual’ method. |
| Switched contraceptives due to pandemic | At risk of unplanned pregnancy (n=1,169) | Reported “I [temporarily/permanently] changed to a different method to prevent pregnancy” in response to “Since the start of the first lockdown, have any of these things happened because of the pandemic?” |
| Accessed services successfully | Tried to access a contraceptive service at least once (n=441) | Reported “Services Accessed during lockdown: Contraception services/advice” and did **not** report “Services tried but failed to access: Contraception services/advice” |
| Faced difficulty accessing services but was able to access in the end | Tried to access a contraceptive service at least once (n=441) | Reported “Services tried but failed to access: Contraception services/advice” and either “I accessed it eventually” or “I used a different service” to question “what happened in the end?” |
| Unable to access contraceptive services | Tried to access a contraceptive service at least once (n=441) | Reported “Services tried but failed to access: Contraception services/advice” and did **not** report “I accessed it eventually” or “I used a different service” to question “what happened in the end?” |
| Successful use of contraceptive services (odds ratio) | All participants (n=1,488) | Reported “Services Accessed during lockdown: Contraception services/advice” |
| Barriers accessing contraceptive services (odds ratio) | All participants (n=1,488) | Reported “Services tried but failed to access: Contraception services/advice” |
| Pregnancy in past year | All participants (n=1,488) | Reported “Yes” to “Are you currently pregnant?” or “In the last year” to “When was most recent pregnancy, even if didn’t carry the baby to term?” |
| London Measure of Unplanned Pregnancy (LMUP) score | Participants with a pregnancy in last 5 years | Questions and scoring detailed at <https://measure.ascody.co.uk/> |
| Unplanned pregnancy in past year | All participants with a pregnancy in past year (n=199) | Reported pregnant in past year and LMUP score <4 |

#### Supplementary Table 2: Service access outcomes – results amongst participants who tried to access contraceptive services (n=441)

| Outcome of attempts to access contraceptive services (% (95% CI)) | | | | | |
| --- | --- | --- | --- | --- | --- |
|  |  | Accessed services successfully | Faced difficulty accessing services but was able to access in the end | Unable to access contraceptive services | Denominators (weighted, unweighted) |
| Total | | 83.6 (79.5, 87.1) | 10.7 (7.9, 14.3) | 5.7 (3.7, 8.6) | 364, 441 |
| Age | | | | | |
|  | 18-24 | 80.8 (70.6, 88.0) | 14.9 (8.7, 24.6) | 4.3 (1.5, 11.7) | 82, 104 |
|  | 25-29 | 83.7 (75.2, 89.7) | 11.7 (6.7, 19.5) | 4.6 (1.9, 10.9) | 104, 136 |
|  | 30-34 | 81.8 (71.1, 89.1) | 9.2 (4.4, 18.5) | 9.0 (4.2, 18.2) | 74, 89 |
|  | 35-44 | 87.1 (79.1, 92.3) | 7.3 (3.6, 14.2) | 5.6 (2.5, 12.2) | 104, 112 |
|  | P-value |  |  |  | p=0.51 |
| Ethnicity | | | | | |
|  | White | 83.7 (79.1, 87.4) | 10.3 (7.4, 14.3) | 6.0 (3.8, 9.3) | 306, 380 |
|  | Black or Black African or Black Caribbean or Black British | 93.4 (66.4, 99.0) | 6.6 (1.0, 33.6) |  | 18, 15 |
|  | Asian or Asian British | 73.4 (48.8, 88.9) | 21.1 (7.8, 45.8) | 5.5 (0.7, 31.4) | 20, 20 |
|  | Mixed or multiple or other ethnic groups | 84.5 (57.4, 95.7) | 8.4 (1.4, 36.2) | 7.1 (1.1, 35.4) | 17, 24 |
|  | P-value |  |  |  | p=0.64 |
| Self-described sexual identity | | | | | |
|  | Heterosexual or Straight | 84.5 (80.3, 88.0) | 9.8 (7.1, 13.4) | 5.7 (3.7, 8.7) | 346, 383 |
|  | Lesbian, Gay, Bisexual or Other | 70.3 (42.8, 88.3) | 22.5 (7.6, 50.7) | 7.1 (1.0, 37.4) | 16, 56 |
|  | P-value |  |  |  | p=0.047 |
| Social Grade | | | | | |
|  | A Upper middle class/ B Middle class | 83.8 (74.5, 90.2) | 13.3 (7.6, 22.2) | 2.9 (0.8, 9.5) | 89, 117 |
|  | C1 Lower middle class/C2 Skilled working class | 85.1 (79.1, 89.5) | 8.7 (5.4, 13.7) | 6.3 (3.5, 10.8) | 187, 211 |
|  | D Working class/ E Lower level of subsistence | 80.4 (70.7, 87.5) | 12.2 (6.8, 21.0) | 7.3 (3.4, 15.1) | 88, 113 |
|  | P-value |  |  |  | p=0.44 |
| Education level | | | | | |
|  | Degree | 85.5 (79.7, 89.8) | 10.7 (7.1, 16.0) | 3.8 (1.8, 7.6) | 193, 234 |
|  | Below degree | 83.1 (76.4, 88.2) | 9.3 (5.7, 15.0) | 7.6 (4.4, 13.0) | 159, 192 |
|  | No qualifications | 60.8 (29.4, 85.2) | 27.6 (8.3, 61.5) | 11.6 (1.7, 49.3) | 12, 15 |
|  | P-value |  |  |  | p=0.13 |
| Living together - relationship but not living together - Single | | | | | |
|  | Single | 87.8 (75.2, 94.4) | 6.5 (2.1, 17.9) | 5.8 (1.8, 17.1) | 50, 61 |
|  | Casual, new, >1, at end or other | 89.1 (70.7, 96.5) | 6.9 (1.6, 25.0) | 4.0 (0.6, 22.5) | 29, 36 |
|  | Married/steady NOT living together | 82.9 (70.0, 90.9) | 11.5 (5.2, 23.5) | 5.7 (1.8, 16.5) | 53, 67 |
|  | Married/steady and living together | 82.2 (76.7, 86.7) | 11.9 (8.3, 16.7) | 5.9 (3.5, 9.8) | 231, 277 |
|  | P-value |  |  |  | p=0.88 |
| Been furloughed under the Coronavirus Job Retention Scheme | | | | | |
|  | No | 83.6 (79.1, 87.4) | 10.5 (7.5, 14.5) | 5.9 (3.7, 9.1) | 308, 373 |
|  | Yes | 85.2 (72.7, 92.5) | 9.9 (4.2, 21.5) | 5.0 (1.5, 15.6) | 54, 65 |
|  | P-value |  |  |  | p=0.95 |
| Became unemployed | | | | | |
|  | No | 83.5 (79.0, 87.2) | 10.5 (7.6, 14.4) | 6.0 (3.9, 9.2) | 322, 390 |
|  | Yes | 87.1 (72.2, 94.6) | 9.4 (3.4, 23.6) | 3.6 (0.7, 17.1) | 40, 48 |
|  | P-value |  |  |  | p=0.79 |
| Number of days drinking | | | | | |
|  | 0 days | 82.2 (74.8, 87.7) | 11.2 (6.9, 17.7) | 6.6 (3.5, 12.2) | 140, 166 |
|  | 1-2 days | 84.0 (76.8, 89.3) | 9.0 (5.2, 15.1) | 7.0 (3.8, 12.8) | 136, 165 |
|  | 3-4 days | 84.2 (72.7, 91.4) | 13.0 (6.6, 24.1) | 2.8 (0.6, 11.7) | 62, 76 |
|  | 5-7 days | 88.4 (68.0, 96.5) | 10.7 (3.1, 31.1) | 0.9 (0.0, 41.4) | 25, 34 |
|  | P-value |  |  |  | p=0.61 |
| Drinking habits compared to pre Covid-19 outbreak | | | | | |
|  | Less these days | 88.8 (81.3, 93.5) | 6.4 (3.0, 12.9) | 4.8 (2.1, 11.0) | 109, 136 |
|  | About the same | 82.3 (75.7, 87.5) | 12.5 (8.2, 18.5) | 5.2 (2.7, 9.9) | 163, 198 |
|  | More these days | 80.5 (70.9, 87.5) | 11.7 (6.5, 20.3) | 7.7 (3.7, 15.5) | 90, 105 |
|  | P-value |  |  |  | p=0.39 |
| Current smoker | | | | | |
|  | No | 87.5 (83.0, 91.0) | 8.3 (5.5, 12.3) | 4.2 (2.3, 7.4) | 269, 327 |
|  | Yes | 72.4 (62.5, 80.6) | 17.4 (11.0, 26.6) | 10.1 (5.4, 18.2) | 94, 113 |
|  | P-value |  |  |  | p=0.0026 |
| Symptoms of depression (PHQ2 score) | | | | | |
|  | No symptoms of depression (0-2) | 85.6 (80.2, 89.7) | 9.3 (6.1, 14.1) | 5.1 (2.8, 9.0) | 212, 259 |
|  | Symptoms of depression (3-6) | 80.5 (73.2, 86.2) | 12.6 (8.1, 19.2) | 6.9 (3.7, 12.4) | 145, 174 |
|  | P-value |  |  |  | p=0.43 |
| Symptoms of anxiety (GAD2 score) | | | | | |
|  | No symptoms of anxiety (0-2) | 87.1 (81.7, 91.1) | 8.2 (5.1, 12.9) | 4.6 (2.4, 8.6) | 202, 235 |
|  | Symptoms of anxiety (3-6) | 78.9 (71.8, 84.6) | 13.9 (9.3, 20.3) | 7.2 (4.0, 12.4) | 159, 203 |
|  | P-value |  |  |  | p=0.11 |

#### Supplementary Table 3: Stopping or switching contraception because of the pandemic – all participants at risk of unplanned pregnancy who used contraception before Covid

| Stopped or switched contraceptive methods (% (95% CI)) | | | | | | | | |
| --- | --- | --- | --- | --- | --- | --- | --- | --- |
|  |  | Stopped using contraceptives | Switched more > less effective | Switched - less > less effective | Switched - more > more effective | Switched less > more effective | Did not switch or stop usual method | Denominators (weighted, unweighted) |
| Total | | 3.6 (2.5, 5.1) | 2.1 (1.3, 3.3) | 4.1 (3.0, 5.8) | 6.6 (5.0, 8.5) | 0.7 (0.3, 1.6) | 82.9 (80.2, 85.4) | 811, 957 |
| Age | | | | | | | | |
|  | 18-24 | 2.6 (0.9, 7.0) | 4.1 (1.8, 9.0) | 10.7 (6.5, 16.9) | 8.5 (4.9, 14.4) | 1.4 (0.4, 5.5) | 72.7 (64.7, 79.4) | 143, 182 |
|  | 25-29 | 5.1 (2.8, 9.1) | 2.7 (1.2, 6.1) | 2.1 (0.8, 5.3) | 6.4 (3.8, 10.6) | 0.4 (0.1, 3.4) | 83.2 (77.5, 87.7) | 209, 261 |
|  | 30-34 | 3.5 (1.6, 7.5) | 1.8 (0.6, 5.4) | 3.4 (1.5, 7.3) | 7.9 (4.7, 12.9) | 1.0 (0.2, 4.3) | 82.4 (76.1, 87.4) | 176, 210 |
|  | 35-44 | 3.0 (1.5, 5.8) | 0.7 (0.2, 2.8) | 2.8 (1.4, 5.6) | 4.9 (2.9, 8.1) | 0.5 (0.1, 2.6) | 88.2 (83.8, 91.5) | 283, 304 |
|  | P-value |  |  |  |  |  |  | p=0.0020 |
| Ethnicity | | | | | | | | |
|  | White | 3.5 (2.4, 5.2) | 1.9 (1.1, 3.3) | 3.1 (2.0, 4.7) | 6.1 (4.6, 8.1) | 0.8 (0.4, 1.9) | 84.5 (81.6, 87.0) | 707, 857 |
|  | Black or Black African or Black Caribbean or Black British | 10.0 (2.8, 30.1) |  | 16.7 (6.3, 37.4) | 13.1 (4.3, 33.5) |  | 60.2 (39.7, 77.6) | 26, 20 |
|  | Asian or Asian British | 2.5 (0.4, 15.4) | 3.4 (0.6, 16.0) | 6.3 (1.9, 19.1) | 5.3 (1.4, 18.0) |  | 82.6 (67.9, 91.4) | 43, 39 |
|  | Mixed or multiple or other ethnic groups | 1.3 (0.0, 25.2) | 5.0 (1.0, 21.9) | 15.4 (6.2, 33.4) | 8.8 (2.6, 26.0) |  | 69.6 (50.8, 83.5) | 31, 39 |
|  | P-value |  |  |  |  |  |  | p=0.0073 |
| Self-described sexual identity | | | | | | | | |
|  | Heterosexual or Straight | 3.5 (2.4, 5.1) | 2.0 (1.2, 3.3) | 4.0 (2.9, 5.7) | 6.3 (4.8, 8.2) | 0.7 (0.3, 1.7) | 83.4 (80.6, 85.9) | 771, 844 |
|  | Lesbian, Gay, Bisexual or Other | 5.9 (1.2, 23.9) | 4.4 (0.7, 22.7) | 4.5 (0.8, 22.8) | 15.0 (5.7, 33.8) | 0.9 (0.0, 34.0) | 69.3 (49.8, 83.7) | 29, 104 |
|  | P-value |  |  |  |  |  |  | p=0.015 |
| Social Grade | | | | | | | | |
|  | A/B - Higher/intermediate managerial, administrative and professional | 2.8 (1.2, 6.2) | 3.3 (1.5, 6.9) | 2.7 (1.1, 6.1) | 8.0 (5.0, 12.7) | 0.4 (0.1, 3.5) | 82.8 (76.9, 87.4) | 200, 251 |
|  | C1/C2 - Supervisory, clerical and junior managerial, administrative and professional, skilled manual workers | 4.4 (2.8, 6.8) | 1.5 (0.7, 3.1) | 3.0 (1.8, 5.1) | 6.2 (4.3, 8.8) | 0.6 (0.2, 1.9) | 84.4 (80.7, 87.5) | 441, 496 |
|  | D/E - Semi-skilled and unskilled manual, casual, lowest grade and unemployed | 2.5 (0.9, 6.3) | 2.1 (0.8, 5.9) | 8.8 (5.4, 14.2) | 5.8 (3.1, 10.6) | 1.6 (0.5, 5.1) | 79.2 (72.4, 84.7) | 169, 210 |
|  | P-value |  |  |  |  |  |  | p=0.020 |
| Education level | | | | | | | | |
|  | Degree | 3.4 (2.1, 5.6) | 1.9 (1.0, 3.7) | 3.5 (2.1, 5.7) | 5.5 (3.7, 8.1) | 1.0 (0.4, 2.5) | 84.6 (81.0, 87.7) | 444, 529 |
|  | Below degree | 3.8 (2.2, 6.4) | 2.3 (1.2, 4.6) | 5.2 (3.3, 8.1) | 7.6 (5.2, 10.9) | 0.2 (0.0, 2.0) | 80.9 (76.4, 84.7) | 349, 408 |
|  | No qualifications | 2.6 (0.1, 39.0) |  |  | 12.9 (3.2, 40.2) | 5.0 (0.5, 35.0) | 79.4 (52.4, 93.1) | 17, 20 |
|  | P-value |  |  |  |  |  |  | p=0.21 |
| Living together - relationship but not living together - Single | | | | | | | | |
|  | Married/steady and living together | 3.8 (2.5, 5.8) | 1.2 (0.5, 2.5) | 3.7 (2.4, 5.6) | 6.1 (4.3, 8.4) | 0.8 (0.3, 2.0) | 84.5 (81.2, 87.3) | 547, 638 |
|  | Married/steady NOT living together | 4.0 (1.6, 9.6) | 3.9 (1.5, 9.5) | 3.9 (1.5, 9.5) | 8.3 (4.4, 15.0) |  | 80.0 (71.6, 86.4) | 115, 142 |
|  | Casual, new, >1, at end or other | 0.7 (0.0, 14.5) | 4.9 (1.5, 14.8) | 10.2 (4.5, 21.4) | 4.5 (1.3, 14.4) | 1.4 (0.2, 11.8) | 78.3 (65.6, 87.3) | 58, 68 |
|  | Single | 3.7 (1.3, 10.3) | 3.3 (1.1, 9.8) | 3.4 (1.1, 10.0) | 8.6 (4.3, 16.5) | 0.9 (0.1, 7.6) | 80.1 (70.6, 87.2) | 91, 109 |
|  | P-value |  |  |  |  |  |  | p=0.20 |
| Been furloughed under the Coronavirus Job Retention Scheme | | | | | | | | |
|  | No | 3.1 (2.0, 4.7) | 2.3 (1.4, 3.7) | 4.2 (2.9, 6.1) | 7.3 (5.5, 9.5) | 0.5 (0.2, 1.5) | 82.7 (79.6, 85.4) | 656, 775 |
|  | Yes | 5.9 (3.1, 11.1) | 1.1 (0.2, 4.9) | 4.0 (1.8, 8.7) | 3.6 (1.6, 8.2) | 1.8 (0.6, 5.9) | 83.5 (76.7, 88.7) | 150, 177 |
|  | P-value |  |  |  |  |  |  | p=0.072 |
| Became unemployed | | | | | | | | |
|  | No | 3.4 (2.3, 4.9) | 2.1 (1.3, 3.5) | 4.0 (2.8, 5.7) | 6.4 (4.8, 8.4) | 0.7 (0.3, 1.7) | 83.4 (80.5, 86.0) | 726, 852 |
|  | Yes | 5.9 (2.4, 13.8) | 1.2 (0.2, 8.5) | 5.7 (2.3, 13.6) | 8.6 (4.1, 17.2) | 1.1 (0.1, 8.6) | 77.5 (67.0, 85.5) | 80, 100 |
|  | P-value |  |  |  |  |  |  | p=0.61 |
| Number of days drinking in past week | | | | | | | | |
|  | 0 days | 4.2 (2.5, 7.0) | 2.4 (1.2, 4.8) | 2.1 (1.0, 4.4) | 6.6 (4.3, 9.9) | 0.3 (0.0, 2.2) | 84.5 (80.1, 88.0) | 321, 375 |
|  | 1-2 days | 3.4 (1.9, 5.9) | 1.6 (0.7, 3.7) | 3.8 (2.2, 6.4) | 7.3 (4.9, 10.6) | 0.8 (0.2, 2.6) | 83.2 (78.8, 86.9) | 334, 399 |
|  | 3-4 days | 2.1 (0.5, 7.5) | 2.5 (0.7, 8.0) | 8.6 (4.5, 15.7) | 5.5 (2.4, 11.8) | 2.5 (0.7, 8.0) | 78.9 (70.1, 85.7) | 107, 121 |
|  | 5-7 days | 4.6 (1.2, 16.5) | 2.1 (0.3, 14.4) | 11.4 (4.8, 24.5) | 3.7 (0.8, 15.5) |  | 78.2 (63.8, 88.0) | 47, 60 |
|  | P-value |  |  |  |  |  |  | p=0.038 |
| Drinking habits compared to pre Covid-19 outbreak | | | | | | | | |
|  | Less these days | 2.9 (1.4, 6.0) | 3.0 (1.5, 6.2) | 3.8 (2.0, 7.1) | 7.5 (4.7, 11.6) | 1.4 (0.5, 4.0) | 81.4 (75.9, 85.9) | 239, 294 |
|  | About the same | 4.2 (2.6, 6.6) | 1.5 (0.7, 3.2) | 3.5 (2.1, 5.8) | 5.8 (4.0, 8.5) | 0.2 (0.0, 1.7) | 84.8 (81.0, 87.9) | 421, 488 |
|  | More these days | 3.1 (1.2, 7.7) | 2.3 (0.8, 6.6) | 6.6 (3.5, 12.0) | 7.5 (4.1, 13.1) | 1.2 (0.3, 5.2) | 79.3 (71.9, 85.2) | 144, 168 |
|  | P-value |  |  |  |  |  |  | p=0.42 |
| Current smoker | | | | | | | | |
|  | No | 3.5 (2.3, 5.3) | 1.8 (1.0, 3.2) | 3.2 (2.1, 4.9) | 6.4 (4.7, 8.6) | 0.8 (0.3, 1.9) | 84.2 (81.2, 86.9) | 636, 752 |
|  | Yes | 3.8 (1.8, 8.0) | 3.0 (1.3, 7.0) | 7.6 (4.4, 12.6) | 7.2 (4.2, 12.2) | 0.5 (0.1, 4.1) | 77.9 (71.0, 83.5) | 173, 203 |
|  | P-value |  |  |  |  |  |  | p=0.11 |
| Symptoms of depression (PHQ2 score) | | | | | | | | |
|  | No symptoms of depression (0-2) | 4.0 (2.6, 6.0) | 1.5 (0.8, 3.0) | 2.5 (1.5, 4.2) | 4.9 (3.4, 7.1) | 0.6 (0.2, 1.7) | 86.5 (83.3, 89.1) | 536, 628 |
|  | Symptoms of depression (3-6) | 2.8 (1.4, 5.7) | 3.2 (1.6, 6.1) | 6.9 (4.4, 10.7) | 9.3 (6.4, 13.5) | 0.8 (0.2, 3.0) | 76.9 (71.4, 81.6) | 266, 319 |
|  | P-value |  |  |  |  |  |  | p=0.00086 |
| Symptoms of anxiety (GAD2 score) | | | | | | | | |
|  | No symptoms of anxiety (0-2) | 3.8 (2.4, 5.9) | 1.8 (0.9, 3.4) | 2.6 (1.5, 4.5) | 6.6 (4.7, 9.1) | 0.8 (0.3, 2.1) | 84.4 (81.0, 87.3) | 507, 584 |
|  | Symptoms of anxiety (3-6) | 3.3 (1.8, 6.1) | 2.3 (1.1, 4.8) | 6.0 (3.8, 9.3) | 6.6 (4.3, 10.1) | 0.7 (0.2, 2.7) | 81.1 (76.2, 85.2) | 298, 367 |
|  | P-value |  |  |  |  |  |  | p=0.26 |

246 respondents (17.4% of total) answered ‘Not applicable’ to questions of usual contraception used as they were already pregnant, planning to get pregnant of unable to get pregnant. 142 respondents (9.5%) answered ‘no method used’ in the year before lockdown. These responses are excluded from the table.
